# Ongoing mitigation strategies and further needs of the United States food industry to control COVID-19 in the work environment

**DOI:** 10.1101/2021.08.06.21261702

**Authors:** Sebastian Llanos-Soto, Ece Bulut, Sarah I. Murphy, Christopher J. Henry, Claire Zoellner, Martin Wiedmann, Diane Wetherington, Aaron Adalja, Samuel D. Alcaine, Renata Ivanek

## Abstract

The COVID-19 pandemic has had dire effects on the United States (US) food industry through impacts on workers’ health and wellbeing and supply chain disruptions. The objectives of this study were to determine what the food industry needs to be able to control COVID-19 impacts in the work environment and what mitigation strategies are being implemented. A web-based needs assessment survey was distributed from January to April 2021, via 13 food professional/trade organizations and 2 social networks, targeting management professionals at food (produce, dairy, poultry, and beef/pork) processing facilities and produce farm operations in the US. Statistical analyses evaluated patterns in self-reported adoption of mitigation strategies against COVID-19 in the participants’ facilities/operations and perceived needs of the industry regarding COVID-Responses to open-ended questions were analyzed using thematic analysis. In total 145 responses were received, of which 79 were usable, including 38 (48%) from the dairy, 17 (22%) from the fresh produce, and 24 (30%) from a mixture of other food industry sectors. Only two usable responses were from the beef/pork sector and none from the poultry sector. Findings revealed that several social distancing, biosecurity, and surveillance mitigation strategies against COVID-19 are commonly implemented in the participants’ facilities/operations, but their implementation frequency differs by the facility/operation size and industry sector. Also, findings indicated that collaboration between the food industry and government agencies, contingency plans and appropriate training, and new technologies are needed to control COVID- 19 in the food industry. Subject to limitations associated with the relatively low response rate (possible selection bias), the findings suggest that the US food industry is prepared to safeguard workers’ health and businesses in the event of a new COVID-19 variant or similar future disaster, provided that appropriate structures are put in place to ensure coordination and compliance, both before and during such an outbreak.

## Introduction

The COVID-19 pandemic, caused by the SARS-CoV-2 beta-coronavirus virus, has presented important challenges to the food industry in the United States (US) and around the world. The food industry has dealt with disruptions in the supply chain [1–5], difficulties meeting market demands and changes in food consumption patterns [2,6,7], negative effects on production capacity [8], labor shortages [9], and decreases in productivity due to absenteeism [10–12], while adopting various public health measures to safeguard its workforce’s health [13]. The US food industry, including transportation and logistics, is more than ever considered critical infrastructure for the nation because of its key role in feeding the US population [14].

The maintenance of such activities has led to the occurrence of COVID-19 outbreaks across all industry sectors in the US, including poultry [15], beef/pork [15], dairy [16], and fresh produce [17]. Indeed, from April 2020 to July 2021, more than 90,000 cases and 450 deaths related to COVID-19 have been reported across food industry sectors in the US [18]. Some of these outbreaks have involved widespread transmission of COVID-19 among employees, as was the case for a central New York State greenhouse in which more than half (171/300) of the workers tested positive for the virus [17]. Food and agriculture workers are amongst the occupations most severely affected by excess deaths due to COVID-19 according to a study comparing the total number of deaths in two scenarios, one theoretical scenario in which COVID-19 never happened (established based on pre-pandemic data for 2018 and 2019) and one scenario that projected deaths during the COVID-19 pandemic [19]. To improve the ability of the US food industry to more effectively and quickly respond to COVID-19 related disturbances and similar future disasters, it is essential to understand the needs and concerns of the food industry regarding the ongoing pandemic. This requires understanding the specific needs and concerns of the various industry sectors, as impacts (e.g., COVID-19 related deaths), challenges, and responses to these challenges might be different across food industry sectors. For instance, industry sectors providing employees with housing and transportation might require additional preventive measures to avoid contact between COVID-19 infected and healthy employees [20].

In the US, governmental institutions, such as the Centers for Disease Control and Prevention (CDC) and the Occupational Safety and Health Administration (OSHA) have published guidelines and checklists to inform and guide the US food industry and other businesses about the correct implementation of social distancing, biosafety, and surveillance strategies to assist in preventing COVID-19 cases [20–23]. The guidelines usually recommend the adoption of multiple methods to socially distance workers, sanitize and clean the workplace and workers’ hands, enforce mask-wearing, and apply surveillance tests to identify COVID-19 cases and prevent further spread. These guidelines were established early in the COVID-19 pandemic (around June 2020) [20, 23] and prior to the widespread availability of vaccines to reduce transmission in the workplace (late 2020 – early 2021) [21, 22]. The proper implementation of these strategies is considered important in reducing the occurrence of COVID-19 cases in the workplace and/or limiting the size of an outbreak; for example, air ventilation and social distancing have been shown to be effective in reducing COVID-19 dissemination among employees in German meat industry facilities in 2020 [24]. Nonetheless, there is a gap in information regarding what the food industry perceives as needed to properly address COVID-19 transmission in food production facilities and operations. Currently, information about the adoption of mitigation strategies in the US food industry remains scarce and limited to certain sectors, such as the meat [15], poultry [15], and dairy sectors [25].

Although, these studies have provided valuable information, they were performed in a comparatively earlier period in the COVID-19 pandemic (April – July, 2020) when the adoption of mitigation strategies by the US food industry could have been different (e.g., vaccine availability) and information, such as guidelines to implement ventilation in buildings [22] and OSHA’s instructions to prevent COVID-19 transmission in the workplace [21], have not yet been published. In addition, the study by Yung et al. [25] was restricted to dairy farmers in the states of Minnesota and Wisconsin, which represents a small part of the US dairy industry and might not reflect the adoption of mitigation strategies in other dairy facilities and operations in the country.

This information is important as knowing how commonly social distancing, biosafety, and surveillance mitigation strategies were adopted to prevent COVID-19 cases across the US food industry and the reasons behind such adoption or lack of adoption are crucial to identify areas where prevention could be improved. Therefore, the objectives of this study were to identify the needs of the US food industry (targeting produce farm operations as well as produce, dairy, poultry, beef/pork processing facilities) in mitigating the impact of the COVID-19 pandemic on workforce and food production, and to assess the ongoing implementation of COVID-19 mitigation strategies in the work environment. These objectives were addressed through the administration of a needs assessment survey between January and April, 2021, detailed in the “Materials and methods” section.

## Materials and methods

### Needs assessment survey design and data collection

A two-part web-based needs assessment survey was developed. Part 1 asked general questions about a survey participant’s overall industry sector (produce, dairy, poultry, beef/pork, or other), and part 2 asked about conditions and COVID-19 mitigation strategies in a food production facility or operation of the participant’s choice (participants who oversee multiple facilities and/or operations were asked to choose one). The wording of questions can be found in S1 Appendix, S1 Table (additionally, the S1 Appendix includes a complete copy of the survey instrument including introduction letter and the consent statement). Briefly, the questions included in part 1 were about a participant’s industry sector, their main role in their organization, how COVID-19 has impacted their industry sector, concerns about COVID-19 control, challenges in maintaining production capacity, needs to successfully mitigate COVID-19, desired features of a computational modeling tool that aids decision making in control of COVID-19, and indicators of a successful response against COVID-19. Part 2 included questions about the industry sector of the participant’s selected facility/operation, maximum tolerable reduction in the production labor force that is compatible with maintaining full production capacity, number and age of employees, availability of employer-provided housing and transportation for employees, importance of different specialized job functions for maintaining production in the event of a COVID-19 outbreak, sources of COVID-19 in the facility/operation, and adoption of COVID-19 mitigation strategies (definitions of mitigation strategies provided to survey participants are shown in Table 1). Part 2 also included several questions applicable only to participants from the produce food sector. Survey questions were initially designed by E.B., R.I., and S.L.-S. The design process included discussing preliminary versions of the survey with A.A., C.Z., D.W., S.D.A., S.I.M., and M.W. until a consensus was reached regarding the phrasing and content of the questions included in the final version of the survey. The survey was piloted between December 8, 2020, and January 6, 2021, by obtaining anonymous responses from 7 out of 10 members of an advisory council for the authors’ COVID-19 research grant comprised of executive-level managers representing the produce, dairy, beef/pork, and poultry industry sectors. Feedback from the piloting process was incorporated into the final version of the survey.

**Table 1.**
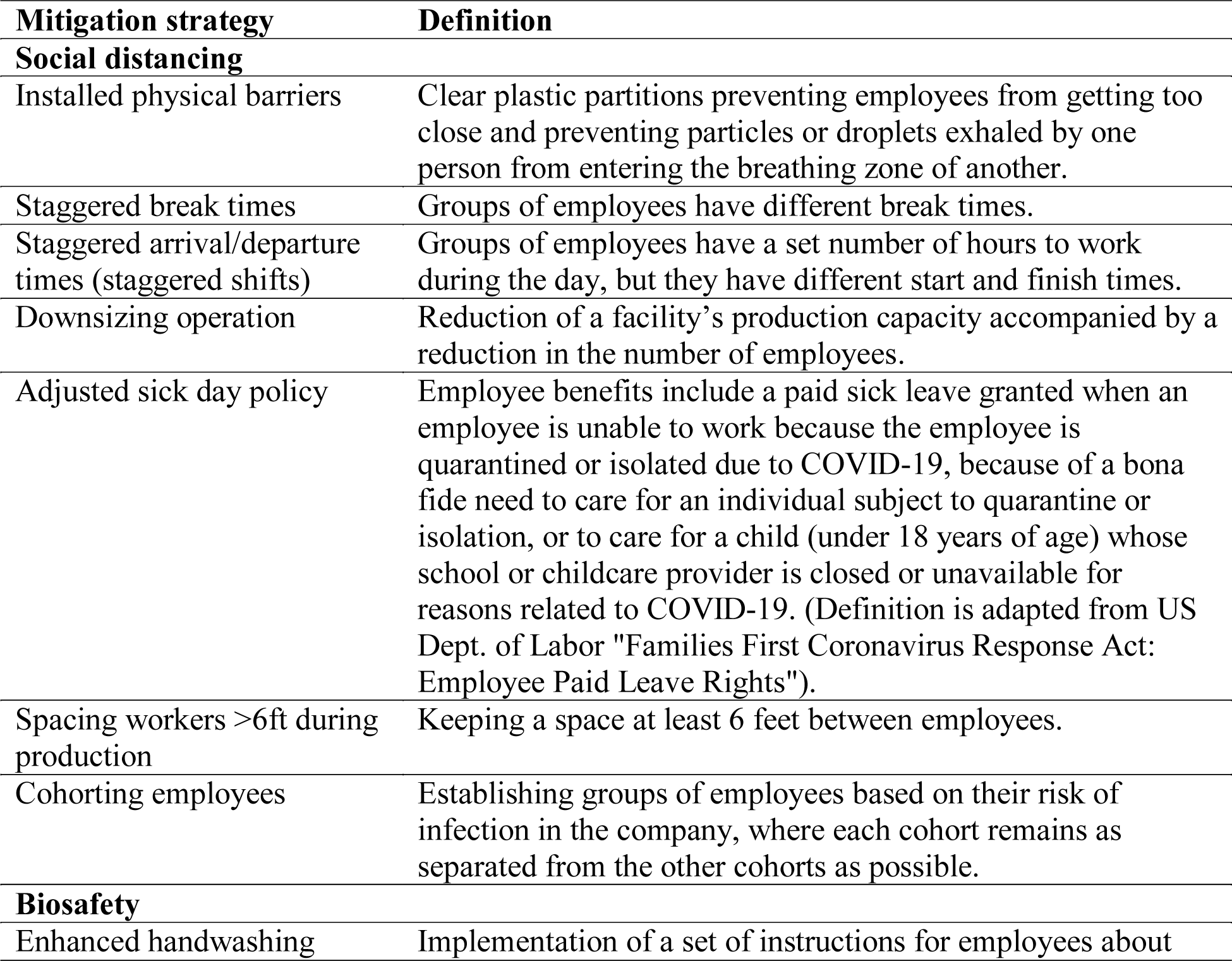
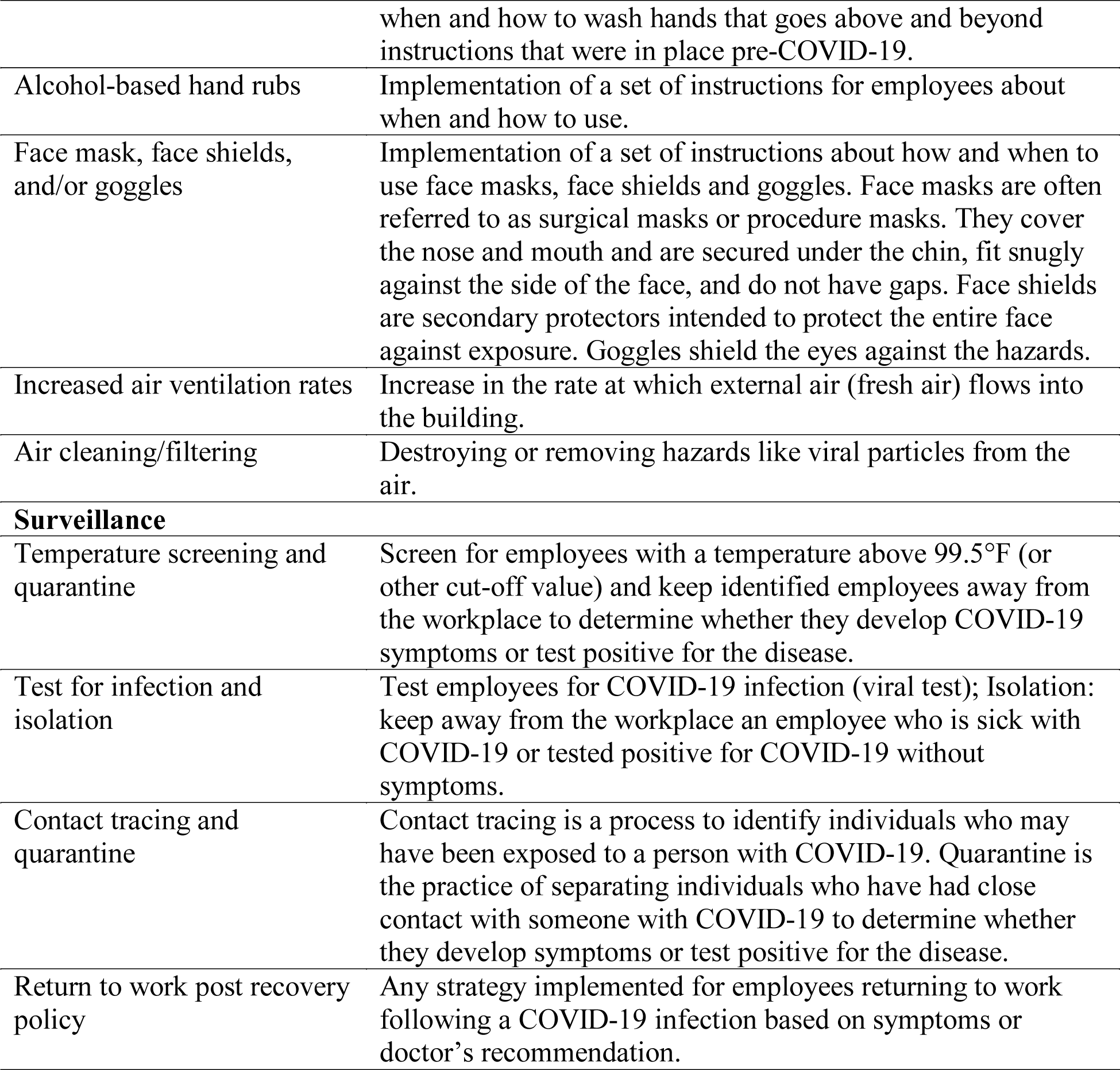
Definitions of social distancing, biosafety, and surveillance mitigation strategies provided to study participants in the needs assessment survey.

The survey was implemented in the Qualtrics survey platform (Qualtrics, Provo, UT) and made available online to survey participants through a link. After clicking on the link participants were shown an introductory letter that provided information about the purpose of the survey and asked to consent to take part in the study. Individuals older than 18 years and affiliated with a produce farm operation or with produce, dairy, beef/pork, poultry, or other food processing facility in the US were considered eligible to participate in the survey; recruitment targeted management professionals at those facilities and operations. No identifying information was collected from participants and their responses remained confidential. No compensation for participation was provided (S1 Appendix). The survey took approximately 30 minutes to complete.

A number of food industry professional/trade organizations were contacted to request their assistance in disseminating the survey to their management professional members. This resulted in successfully collected responses via 13 organizations, including 3 associated with the fresh produce industry, 5 with dairy, 1 with beef/pork & poultry, and 4 with general processing. Additionally, the survey was distributed via two social networks of study authors (Table 2). The period for survey distribution started on January 19, 2021, and concluded on April 6, 2021. The study was approved by the Cornell University Institutional Review Board for Human Participants (IRB protocol #2006009660). No power-based sample calculation was conducted due to the preliminary nature of the investigation.

**Table 2.**
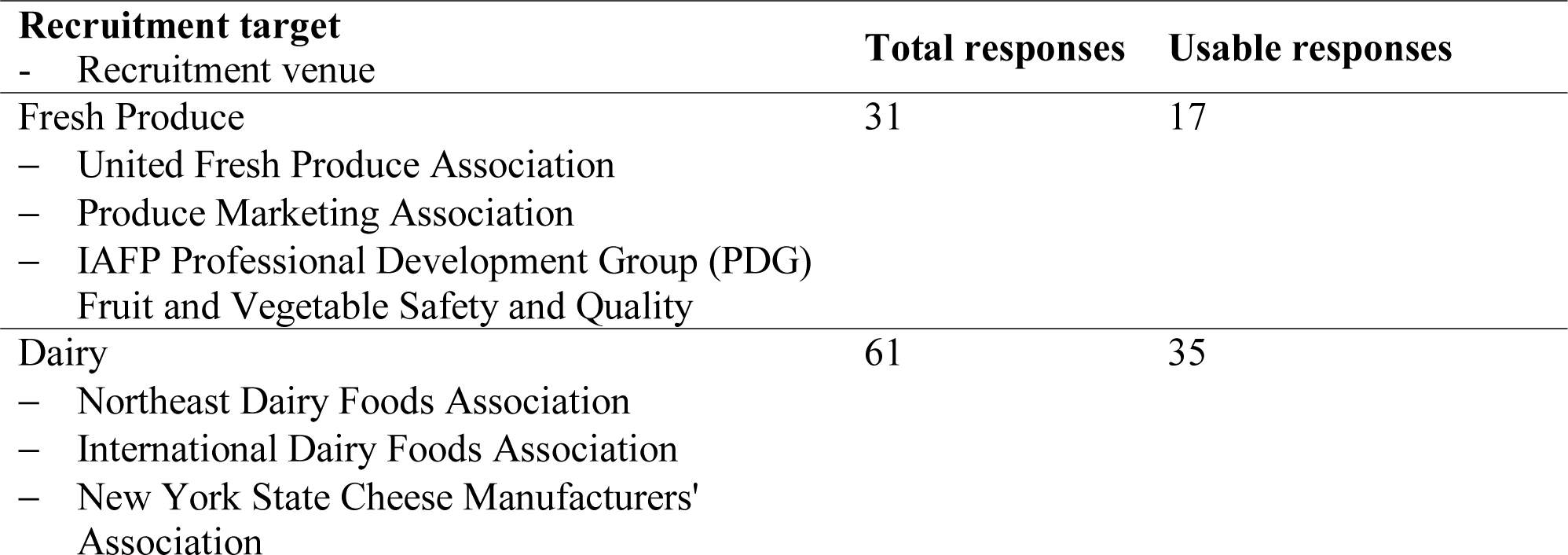
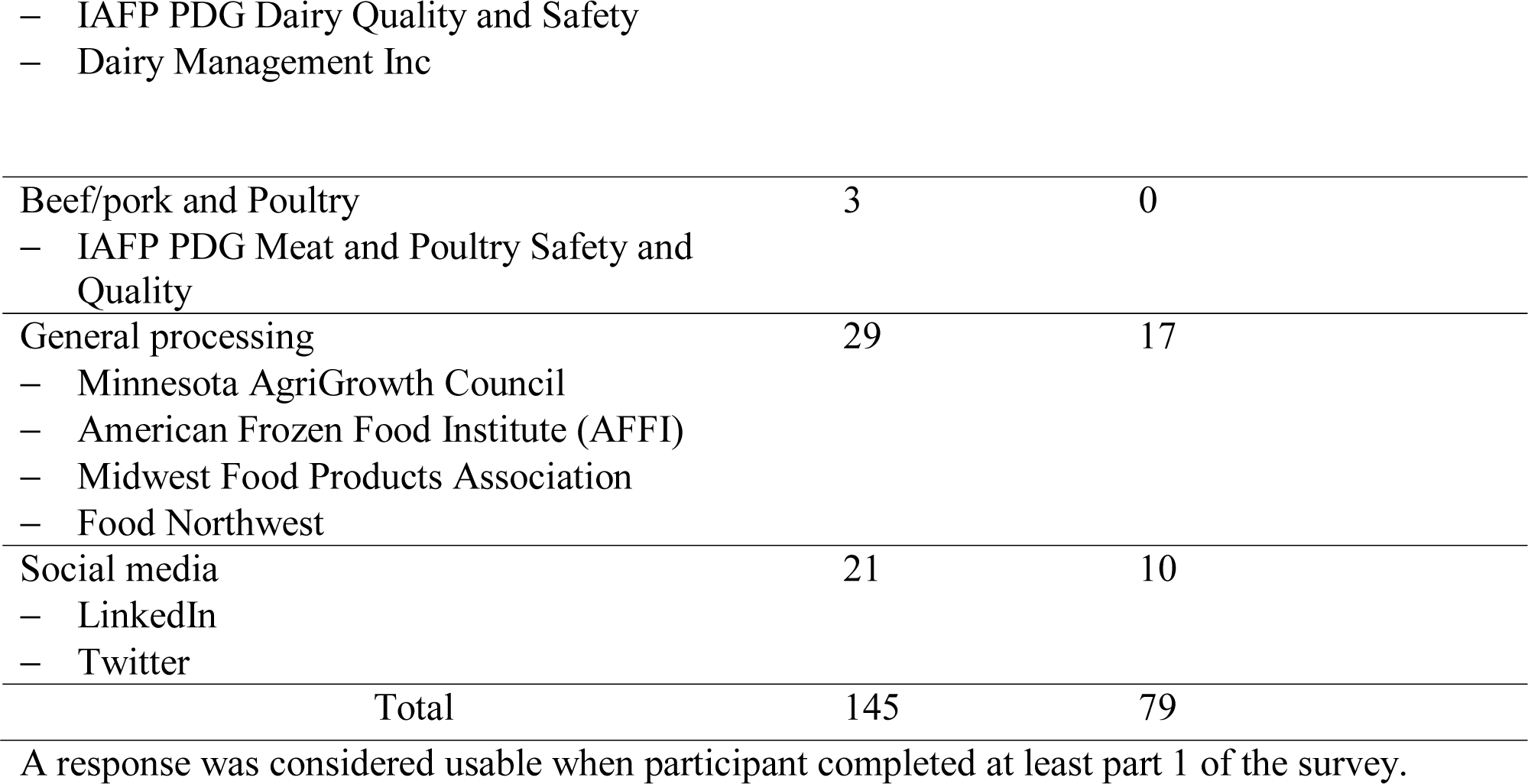
Number of needs assessment survey responses (total received and usable) by the industry sector and social media recruitment targets.

### Data management and statistical analysis

Only responses from participants that completed at least part 1 of the survey were considered usable (Table 2). These were organized in an Excel datasheet (Microsoft, Seattle, WA) for subsequent analysis. Responses to multiple choice and open-ended questions asking for specific numbers were treated as numerical data for statistical analysis. If a participant responded an interval to a question asking for a specific number , then the mean of the interval was used.

Incomplete responses or those unrelated to the corresponding questions were included as missing values in the datasheet. Statistical analysis was carried out in R v. 4.0.3 [26]. Responses to the survey questions were summarized and organized in tables and visualized using heatmaps.

Responses to numeric open-ended questions were summarized using median, mean, interquartile range (IQR), and range. Levels in Likert items were treated as interval (with assigned values values 1-5) or numerical (with assigned values 0, 0.5 or 1) data for statistical analysis, so that the median score of responses to a Likert item could be calculated. Responses to the question Q20 “What was the average number of employees in this facility/operation in 2019?” were transformed from interval to nominal data by grouping responses in the levels “Small” (1-49 employees), “Medium” (50-249 employees), and “Large” (>250 employees) based on criteria established by the Organisation for Economic Co-operation and Development (OECD) [31]; this variable served as a proxy for facility/operation size.

A complete plan of analysis of survey questions can be found in S1 Appendix, S1 Table, including questions identified as the outcomes of interest and independent variables (predictors) in the analysis of associations. As applicable, Kruskal-Wallis, Fisher’s exact, Mann-Whitney U or Spearman’s rank correlation tests were used to identify predictors associated with the outcomes of interest at the bivariable level. Associations were considered significant at *p* ≤ 0.05.Post-hoc assessment of significant associations was carried out using the Dunn’s test to determine specific statistical differences between variable levels. Obtained *p*-values were adjusted for multiple testing using the false discovery rate (FDR) [27]. The data underlying the results presented in the study are available in Zenodo (doi: 10.5281/zenodo.5165334).

### Thematic analysis

Thematic analysis involves determining common themes or ideas that are repeated across participants’ open-ended responses to a certain question [28]. As a follow-up to certain Likert questions or multiple-choice questions, we asked an open-ended question to gain a deeper understanding of participants’ opinions/perceptions. These open-ended questions were preliminarily assessed to select questions with diverse and informative responses for thematic analysis. In other words, questions with responses that provided new information (i.e., not being accounted for in responses to other questions in the survey) and diverse enough to generate at least two codes were selected. During the initial assessment, a preliminary codebook was generated to classify participants’ responses into codes and subsequently identify themes emerging from those codes. This codebook was further refined through discussions among authors (S.L.-S., S.I.M., E.B., and R.I.). Theme identification across questions was first carried out individually by authors (S.L.-S., S.I.M., E.B., and R.I.), and a final consensus was reached following discussion. Details for codes and themes can be found in S2 Table.

## Results

### Responses from 79 survey participants could be analyzed, most being from the dairy industry sector and acting as corporate food safety and quality managers

In total, 145 survey responses were collected. However, based on the decision to only include responses from participants that completed at least part 1 of the survey (usable responses), responses from only 79 participants were retained for statistical (Tables 3-5) and thematic analysis (Table 6). The following results were obtained from responses to questions in part 1, where participants answered general questions about the industry sector to which they belong (Table 3). Among 79 participants, 38 (48%) were from the dairy industry sector, 17 (22%) from fresh produce, and 22 (28%) from other food industry sectors (e.g., chocolate production, frozen food, prepared food, wine production, cereals) (Q1, Table 3). Only two responses were obtained from the beef/pork industry and thus were grouped into the “Other” category for statistical analysis. Three participants self-reported association with all 4 food industry sectors (i.e., Fresh produce, Dairy, Poultry, and Beef/pork) and thus were grouped in the “Other” category. No responses were received from the poultry sector. (Note: as stated, Q1 in part 1 asked for the industry sector of a participant, which is different from Q15 in part 2 that asked about the industry sector of a facility/operation the participant chose to describe; this is why responses to Q1 and Q15 are slightly different although they both ask about the industry sector.)

**Table 3.** Number and proportion of responses among survey participants (N=79) for questions, analyzed as categorical variables, from each of the two parts of the needs assessment survey (part 1: General questions about a participant’s industry sector; part 2: Conditions and COVID-19 controls in a facility or operation of participant’s choice).

| Q<sup>a</sup> | Variable | Response | Number | % |
| --- | --- | --- | --- | --- |
| **Part 1** |  |  |  |  |
| Q1<sup>b</sup> | What industry sector are you in?<br>(select all that apply) | Fresh produce | 17 | 22 |
|  |  | Dairy | 38 | 48 |
|  |  | Poultry | 0 | 0 |
|  |  | Beef/pork<sup>c</sup> | 2 | 3 |
|  |  | Other<sup>d</sup> | 22 | 28 |
| Q2<sup>b</sup> | Select your main role within your organization | C-suite | 13 | 16 |
|  |  | Regional manager | 0 | 0 |
|  |  | Facility manager | 21 | 26 |
|  |  | Research and development | 3 | 4 |
|  |  | Corporate food safety and quality | 29 | 37 |
|  |  | Other<sup>e</sup> | 11 | 14 |
|  |  | Prefer not to answer | 2 | 3 |
| Q3 | Did COVID-19 have a significant impact on your industry sector? | Yes | 67 | 85 |
|  |  | No | 12 | 15 |
| Q4 | In which way(s) has COVID-19 significantly impacted your industry sector? (select all that apply) | Operations/production has been reduced/cut back | 38 | 57 |
|  |  | Operations/production has expanded | 23 | 34 |
|  |  | Implemented robotics, sensors, automation, and/or computer modeling | 5 | 7 |
|  |  | Management/corporate employees working remotely | 52 | 78 |
|  |  | Major changes in operational staffing control and protection protocols | 54 | 81 |
|  |  | Other | 1 | 1 |
| **Part 2: Conditions and COVID-19 controls in a facility or operation of participant's choice** |  |  |  |  |
| Q15<sup>b</sup> | In what industry sector is this facility/operation? | Fresh produce | 17 | 22 |
|  |  | Dairy | 40 | 51 |
|  |  | Poultry | 0 | 0 |
|  |  | Beef/pork | 1 | 1 |
|  |  | Other<sup>f</sup> | 20 | 25 |
|  |  | No response | 1 | 1 |
| Q20<sup>g</sup> | What was the average number of employees in this facility/operation in 2019? | Small (1-49 workers) | 25 | 32 |
|  |  | Medium (50-249 workers) | 33 | 42 |
|  |  | Large (>250 workers) | 16 | 20 |
|  |  | Prefer not to answer | 1 | 1 |
|  |  | Missing | 4 | 5 |
| Q22 | Does this facility/operation provide group temporary (seasonal) housing to any of your employees? | Yes | 9 | 11 |
|  |  | No | 67 | 85 |
|  |  | No response | 3 | 4 |
| Q24 | Does this facility/operation provide group transportation services (bus, truck, etc.) to employees to/from work? | Yes | 8 | 10 |
|  |  | No | 68 | 86 |
|  |  | No response | 3 | 4 |
| Q31 | If in the future the available labor force in this facility/operation is affected by a COVID-19 outbreak, what short/mid-term solutions should be considered to maintain production? (check all that apply) | Extend the number of work hours for remaining workers | 59 | 75 |
|  |  | Backfill with emergency personnel from third-party companies | 28 | 35 |
|  |  | Backfill by reorganizing personnel in the same facility | 48 | 61 |
|  |  | No response | 7 | 9 |
| Q32 | To prevent labor shortage in this facility/operation due to a potential future COVID-19 outbreak, should capital investment into mechanization be considered as a long-term solution to maintain production? | Yes | 57 | 72 |
|  |  | No | 13 | 16 |
|  |  | No response | 9 | 11 |
| Q43<sup>b</sup> | What was the main reason for your choice to describe conditions and COVID-19 mitigation in this particular food production facility/operation? | I am mostly familiar with this facility/operation | 24 | 30 |
|  |  | This is our unique facility/operation | 5 | 6 |
|  |  | This is our typical facility/operation | 16 | 20 |
|  |  | This facility/operation has been impacted greatly by COVID-19 | 11 | 14 |
|  |  | This is our strategically important facility/operation | 13 | 16 |
|  |  | Other | 4 | 5 |
|  |  | No response | 0 | 0 |
| *Questions applicable only to the fresh produce industry sector* |  |  |  |  |
| Q16<sup>h</sup> | How does this facility/operation operate? | Year-round | 15 | 88 |
|  |  | Seasonally | 2 | 12 |
|  |  | No response | 0 | 0 |
| Q18<sup>h</sup> | What role best describes this facility/operation? | Grower | 2 | 12 |
|  |  | Packing House | 2 | 12 |
|  |  | Processor | 4 | 24 |
|  |  | Grower and Field packer | 5 | 29 |
|  |  | Grower and Processor | 3 | 18 |
|  |  | Other | 1 | 6 |
|  |  | No response | 0 | 0 |
| Q19<sup>i</sup> | Please select which part of your Grower and Processor operation will you describe in the remaining questions? | Grower operation | 0 | 0 |
|  |  | Processor facility | 3 | 100 |
|  |  | No response | 0 | 0 |

<sup>a</sup> Question (Q) number in the needs assessment survey.
<sup>b</sup> Participants were required to answer the question to continue the needs assessment survey.
<sup>c</sup> Two participants from the beef/pork industry were grouped in the “Other” category for statistical analysis.
<sup>d</sup> Participants’ provided responses for the “Other” industry sector (number responses): Food processing (2); Frozen fruits (2); Chocolate (1); Cereal (1); Co-packaging of shelf-stable products (1); Consumer packaged goods with fresh produce (1); Food manufacturing (1); Food service (1); Frozen produce (1); Manufacturing shelf-stable foods (1); Prepared food (1); Restaurants (1); Seafood (1); Spirits (1); Sugar (1); Vegetable processing (1). Additional 4 participants were grouped into the “Other” category for statistical analysis: 1 participant who is an academic affiliated with the food industry and 3 participants who reported affiliation with all 4 sectors (i.e., Fresh produce, Dairy, Poultry, and Beef/pork).
<sup>e</sup> Participants’ provided responses for the “Other” main role within their organization (number responses): Quality assurance manager (3); Sales service (1); Affineur (1); Human resources director (1); Owner (1); Office manager (1); CEO (1); Emeritus professor (1); Grower (1).
<sup>f</sup> Participants’ provided responses grouped under the “Other” industry sector for the described facility/operation of the participant’s choice (number responses): Cleaning and sanitation (1); Restaurants (1); Chocolate (1); Prepared food (1); Food manufacturing (1); Shelf-stable products (2); Frozen Produce (1); CPG sauces (2); Cereal (1); Food processing (2); Beverage, spirits (1); Vegetable processing (1); Frozen fruits (2); Seafood (1); Sugar (1). Additionally, one participant who answered “Other” specified working in all industry sectors.
<sup>g</sup> Original levels “Less than 10”, “10-49”, “50-99”, “100-249”, “250-499”, “500-999”, “1000-2000”, and “More than 2000” were grouped in the levels “Small” (1-49 employees), “Medium” (50-249 employees), and “Large” (>250 employees) based on criteria established by the Organisation for Economic Co-operation and Development (OECD).
<sup>h</sup> This question was only shown to 17 participants who selected the option “Fresh produce” in question Q15.
<sup>i</sup> This question was only shown to 3 participants who selected the option “Grower and Processor” in question Q18.

**Table 4.** Summary statistics for questions included in part 2 of the needs assessment survey (that asked about conditions and COVID-19 controls in a food production facility or operation of participant’s choice), which were analyzed as interval variables (number of survey participants =79).

| Q<sup>a</sup> | Variable | Number of responses | Mean | Median | IQR | Range | No response |
| --- | --- | --- | --- | --- | --- | --- | --- |
| Q26 | What is the largest percent reduction in the general production labor force that this facility/operation could withstand over a period of one week without reduction in the production capacity? | 66 | 15 | 15 | 10-15 | 5-50<sup>b</sup> | 13<sup>c</sup> |
| Q21 | What is the approximate proportion (%) of employees in this facility/operation that are between 50-69 years of age?<sup>d</sup> | 66 | 32 | 28 | 20-40 | 0-100 | 13 |
| Q21 | What is the approximate proportion (%) of employees in this facility/operation that are 70 years old or older?<sup>d</sup> | 62 | 3 | 1 | 0-5 | 0-17 | 17 |
| Q23 | Approximately what proportion (%) of employees in this facility/operation are provided with group temporary housing?<sup>e</sup> | 7 | 49 | 35 | 35-70 | 10-100 | 2 |
| Q25 | Approximately what proportion (%) of employees in this facility/operation are provided with group transportation to/from work?<sup>f</sup> | 7 | 40 | 28 | 15-63 | 10-90 | 1 |

<sup>a</sup> Question (Q) number in the needs assessment survey
<sup>b</sup> This was a single-choice question with available responses: 5%, 10% , 15%, 20%, 30%, 40%, 50%, and Do not know; thus, the observed range spanned the full range of possible responses.
<sup>c</sup> Nine participants responded "Do not know" and were grouped with "No response" for analysis.
<sup>d</sup> The original question asked about '50-69 years of age' and '70 years or older' but were separated here to more clearly indicate their values.
<sup>e</sup> This question was only shown to 9 participants who selected "Yes" in question Q22: "Does this facility/operation provide group temporary (seasonal) housing to any of your employees?".
<sup>f</sup> This question was only shown to 8 participants who selected "Yes" in question Q24: "Does this facility/operation provide group transportation services (bus, truck, etc.) to employees to/from work?".

**Table 5.**
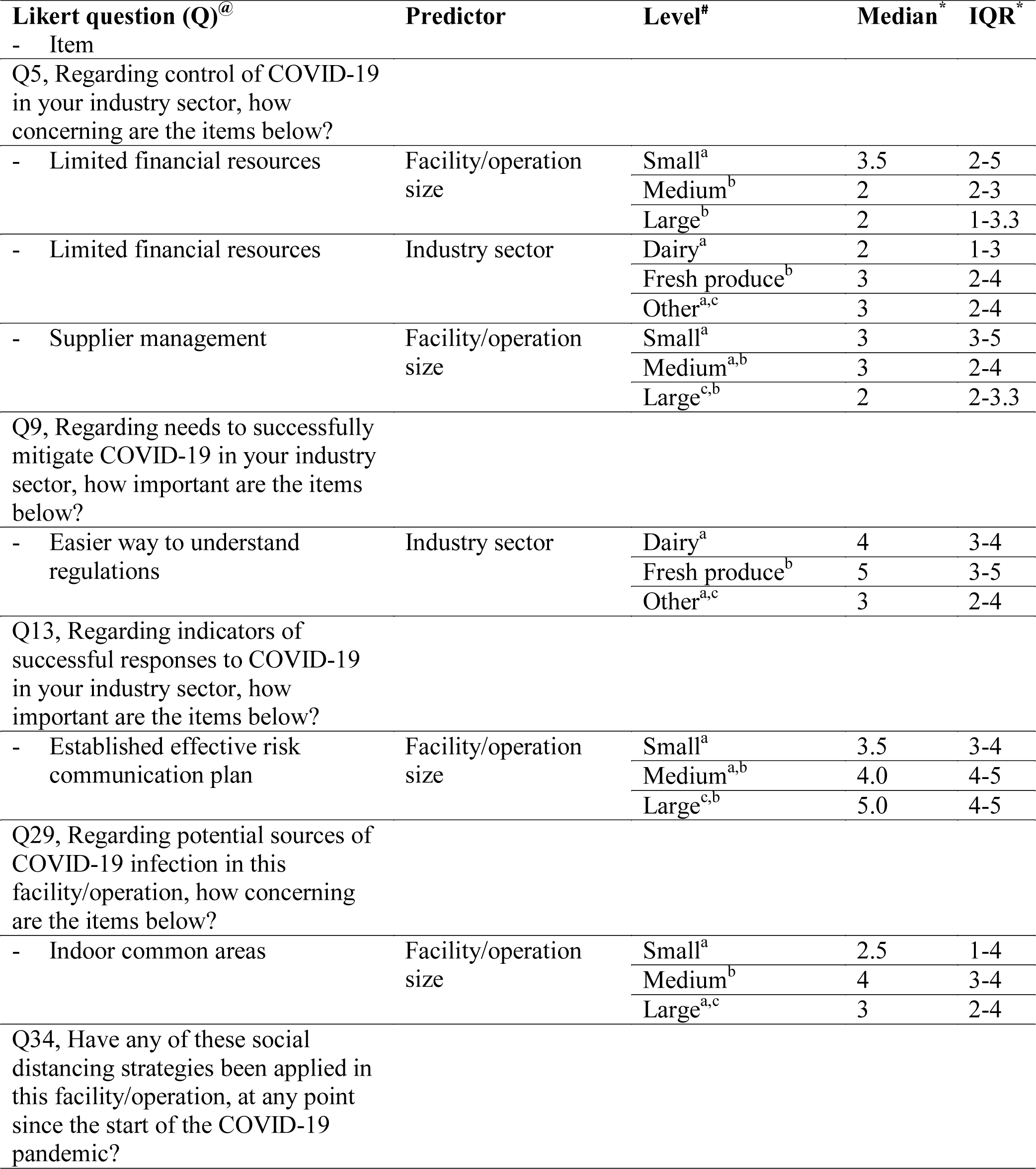
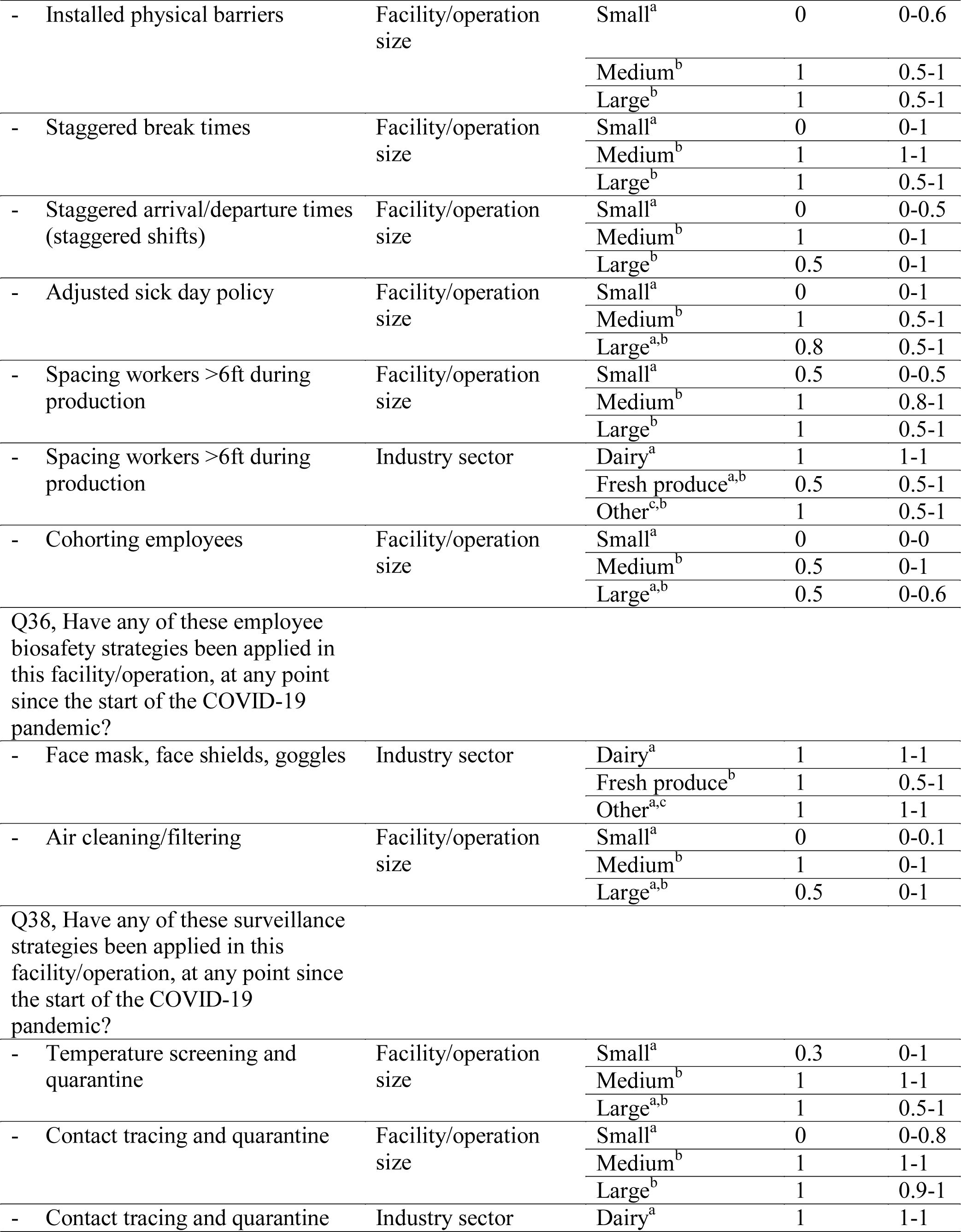
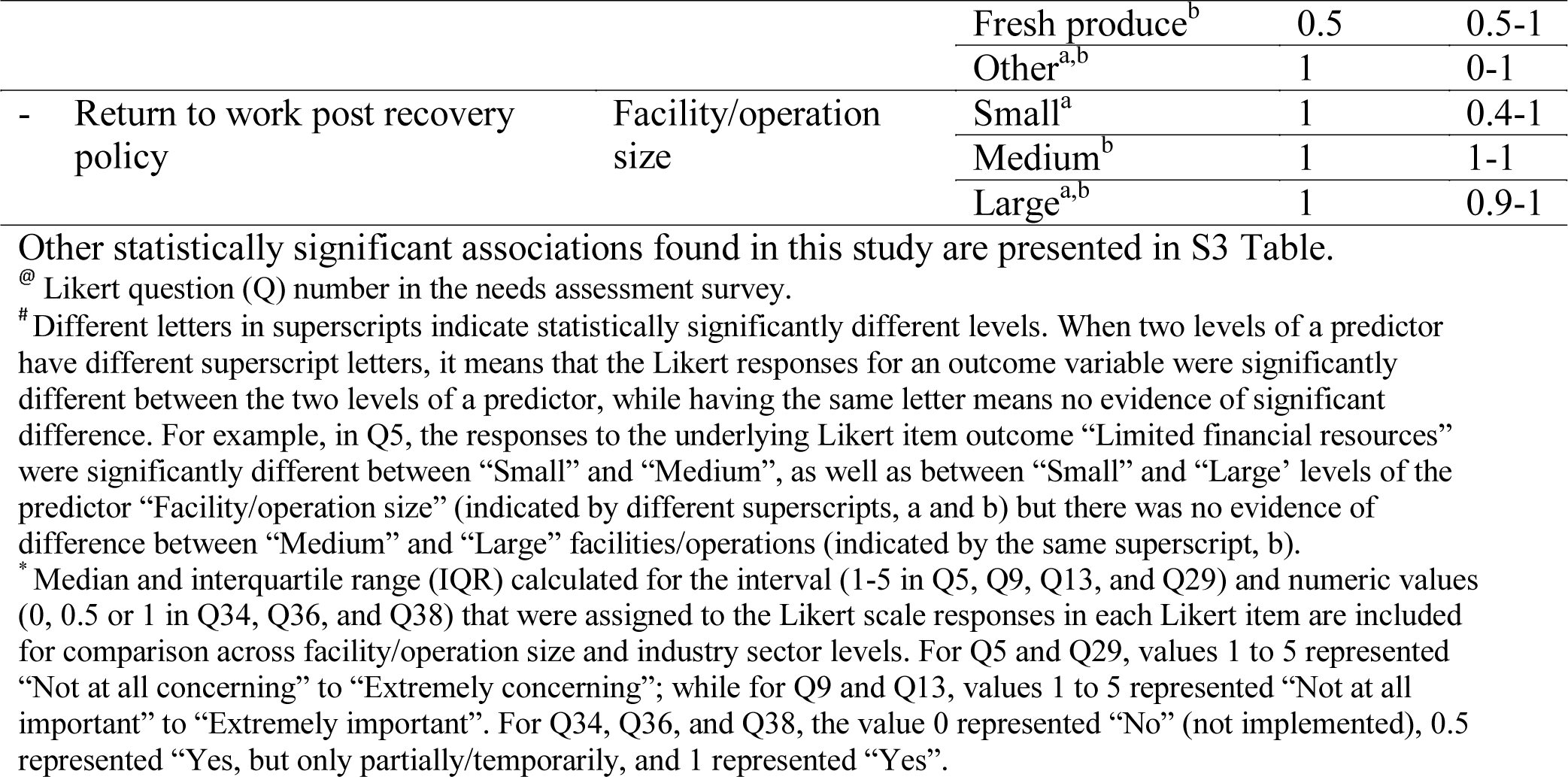
Significant associations found in the bivariable analyses (*p*≤ 0.05) between a specific Likert item (outcome) in a survey question (Q) and independent variables (predictors) describing “Industry sector” (Q1) and “Facility/operation size” (Q20) after post-hoc analysis and false discovery rate adjustment.

**Table 6.**
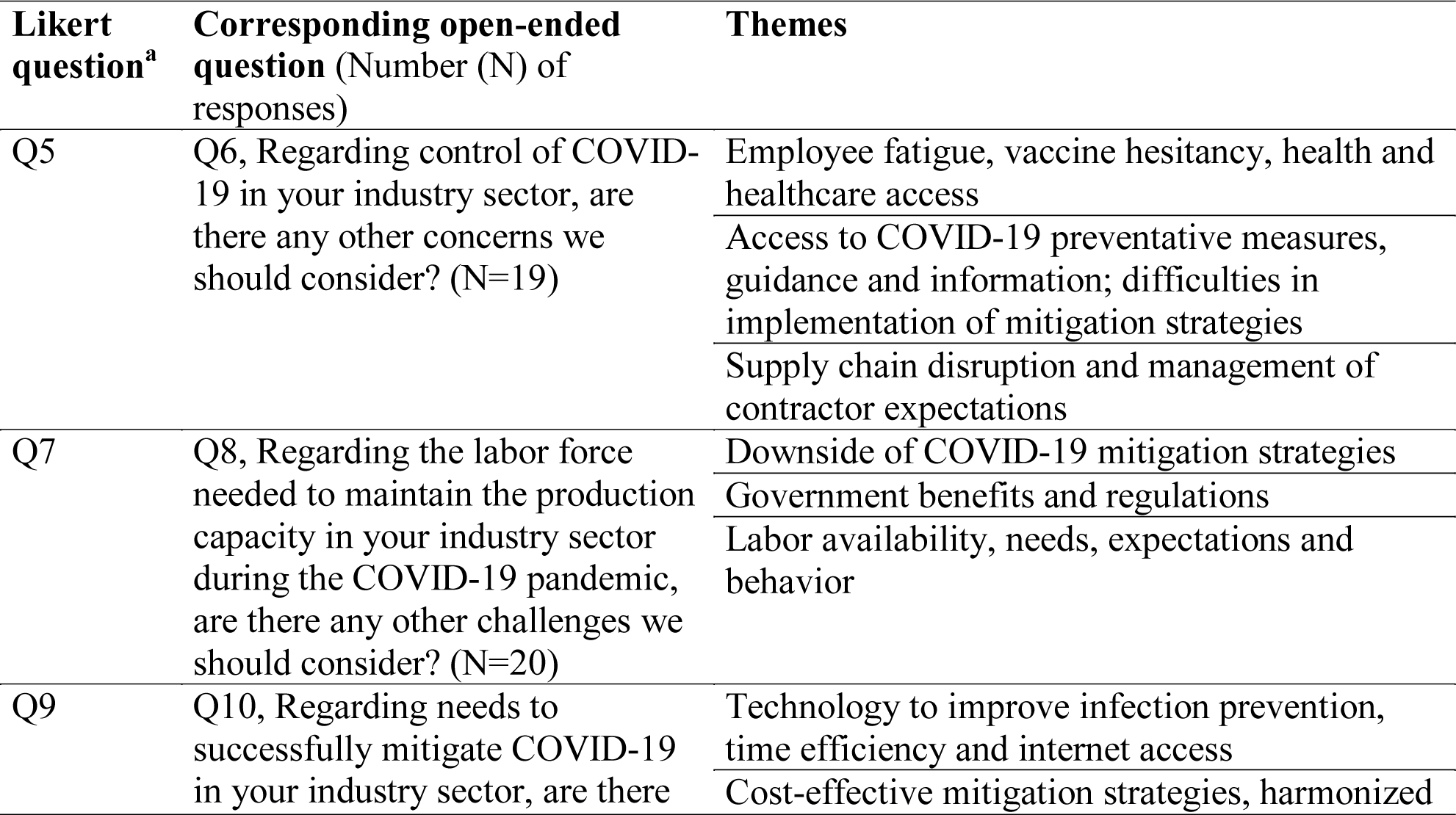
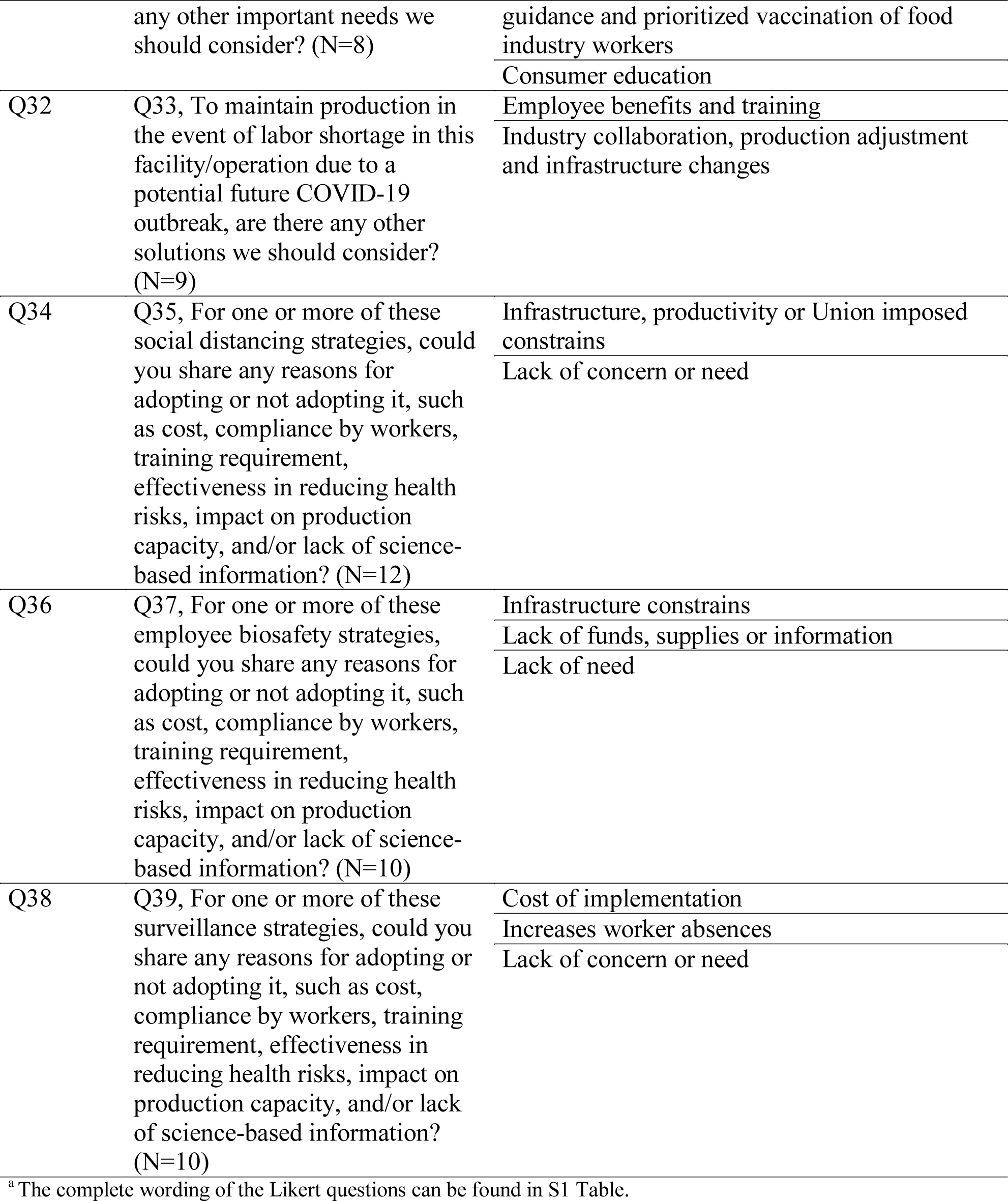
Themes identified in participants’ responses to selected open-ended questions in the needs assessment survey, the purpose of which was to provide depth to the corresponding Likert items questions.

The most common roles of the 79 participants in their organizations were corporate food safety and quality manager (29, 37%), followed by facility managers (21, 27%), and c-suite executive (13, 16%) among others (Q2, Table 3). Most (67/79, 85%) indicated that COVID-19 has significantly impacted their industry sector. The most common ways COVID-19 has significantly impacted their industry sector was through major changes in operational staffing control and protection protocols (54/79, 68%), remote work of management or corporate employees (52/79, 66%), and reduced or cut back operations/production (38/79, 48%) (Q4, Table 3).

### Participants’ chosen facilities/operations could withstand up to a median of 15% reduction of the production labor force over a week while still being able to maintain the full production capacity

The following results include responses from part 2 of the needs assessment, which asked about a facility or operation of participant’s choice (Tables 3 and 4). The most common reason for selecting a facility to describe in the survey was participant’s familiarity with the facility (24/79, 30%) (Q43, Table 3). In terms of the average number of employees in 2019, most participants (33/79, 42%) chose to describe a medium size-facility/operation (50-249 employees) (Q20, Table 3). The largest percent reduction in the general production labor force over a week that the participants’ facility/operation could withstand without reducing their production capacity had a median of 15% (IQR: 10%-15%; Q26, Table 4). Only 11% and 10% of the participants’ facilities/operations provided group temporary (seasonal) housing and transportation to employees, respectively (Q22 and Q24, Table 4). Regarding short/mid-term solutions to maintain production and prevent their facility/operation labor force from being affected in the event of a COVID-19 outbreak, most of the participants’ facilities and operations considered extending the number of work hours for remaining workers (59/79, 75%) and/or backfilling by reorganizing personnel in the same facility (48/79, 61%) (Q31, Table 3). Most participants (57/79, 72%) agreed that capital investment into mechanization should be considered as a long-term solution to maintain production in the face of a future COVID-19-associated labor shortage (Q32, Table 3). Further, participants suggested employee- and production schedule adjustments as additional solutions to maintain production (thematic analysis of Q33, Table 6).

### Ever-changing government regulations and easier ways to understand these regulations as well as establishing risk communication plans were deemed important to participants for successful mitigation of COVID-19 in their industry sector

Regarding the control of COVID-19 in their industry sector (Q5, Fig 1A), *Labor availability* and *Complex ever-changing government regulations* were considered very concerning (median score = 4) among the majority of participants. Conversely, *Workers’ abuse of control measures*, *Limited financial resources*, and *Product quality* were only slightly concerning (median score ≤ ), according to most survey participants (Fig 1A). Kruskal Wallis tests showed that the reported concern for *Limited financial resources* was significantly higher in small compared to medium (*p* = 0.04) and large-sized facilities/operations (*p* = 0.04) and was also significantly higher in the fresh produce industry sector compared to dairy (Q5, *p* = 0.05; Table 5). Similarly, the reported concern for *Supplier management* was also significantly higher in small compared to large-sized facilities/operations (Q5, *p* = 0.03; Table 5). In response to the open-ended question about additional concerns regarding the control of COVID-19 in their industry sector, participants stated concerns related to employees, COVID-19 mitigation, and supply chain factors (thematic analysis of Q6, Table 6). Regarding employee’s mental health, a participant stated the following: “Mental health impact on managerial and office staff. In talking with industry colleagues, my experience is that many facilities have cut back on the staff responsible for ensuring that the facility is operating in an efficient and structured manner, while expanding the production capacity of the facility. It leaves many of these employees in a position where they are stretched thin and feel overwhelmed.” Participant ID #75, Other (co-packaging of shelf stable products) Among challenges associated with the labor force needed to maintain the production capacity (Q7, Fig 1B), most participants perceived that all of the items presented to them were at least moderately challenging (median score ≥ ), with the *Need to train labor*, *Access to workers with necessary skills*, and *Access to number of workers needed* being considered very challenging (median score = 4). Additional challenges to maintaining the production capacity were related to the COVID-19 mitigation, employee and government factors (thematic analysis of Q8, Table 6). In particular, one participant expressed their frustration when dealing with continuously changing government regulations:

**Fig 1.**
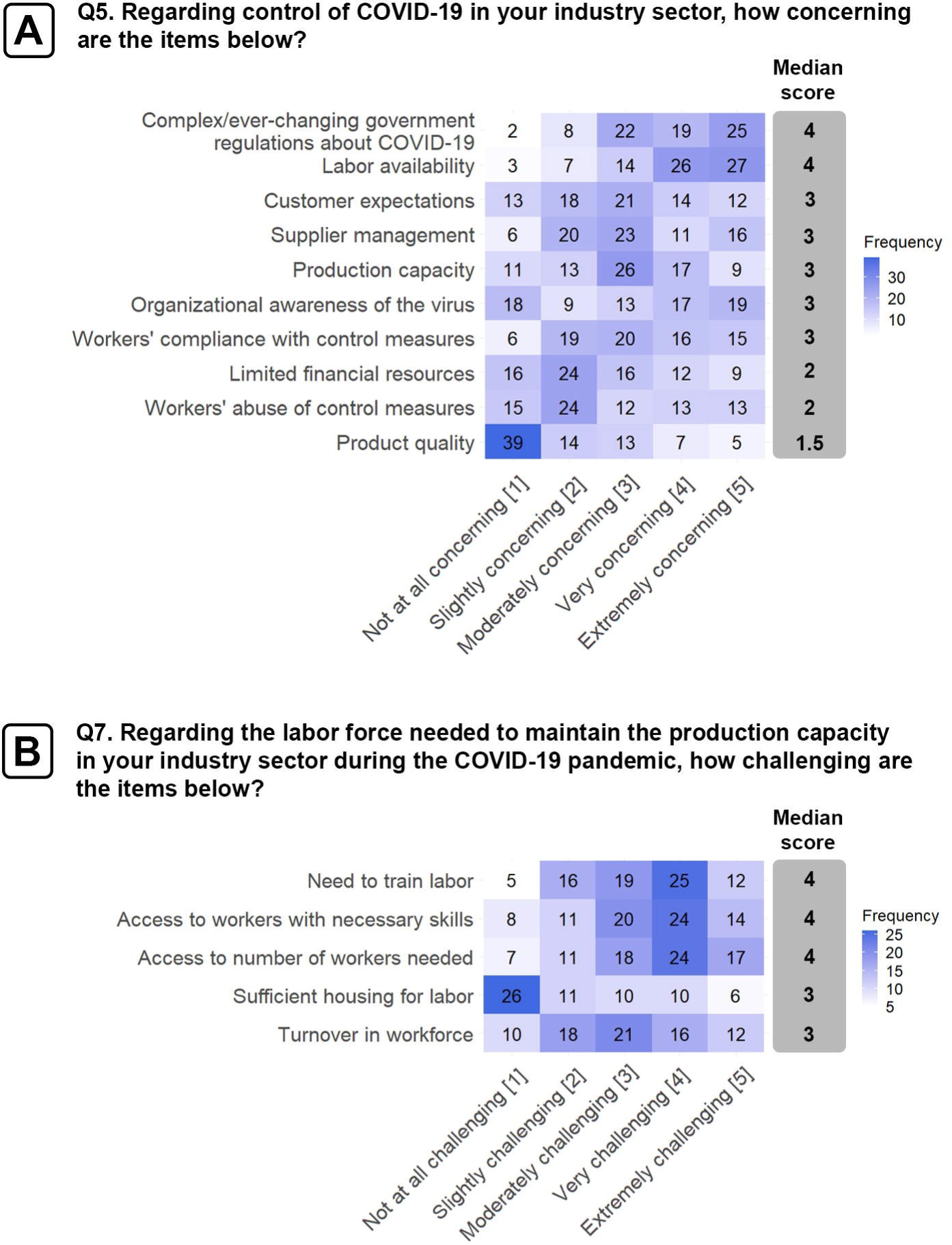
Heatmaps representing responses to 5-point scale Likert questions regarding [A] concerns about COVID-19 control and [B] challenges to maintain production. These Likert questions were included in part 1 of the needs assessment survey asking general questions about a participant’s industry sector. For each item within a Likert question, a median shown to the right of the heatmap was calculated from the interval values (1-5), assigned to answers ranging from “Not at all concerning” to “Extremely concerning” [A] and “Not at all challenging” to “Extremely challenging” [B].

“Government directives and laws regarding labor, time off, and pay rules and the changes in these made managing government special rules a full-time job, rather than managing the pandemic we were managing the labor law compliance tasks.” Participant ID #45, Fresh produce.

In terms of the needs to successfully mitigate COVID-19 in their industry sector (Q9, Fig 2A), most participants indicated that *Easier way to understand regulations* was the most important (median score = 3.5), being valued as a more important need by the fresh produce industry compared to the dairy industry sector and the group of “Other” industry sectors (*p* = 0.05 and *p* = 0.03, respectively; Table 5). In the corresponding open-ended question participants further stated needs for improved technology solutions, mitigation strategies and consumer education (thematic analysis of Q10, Table 6).

**Fig 2.**
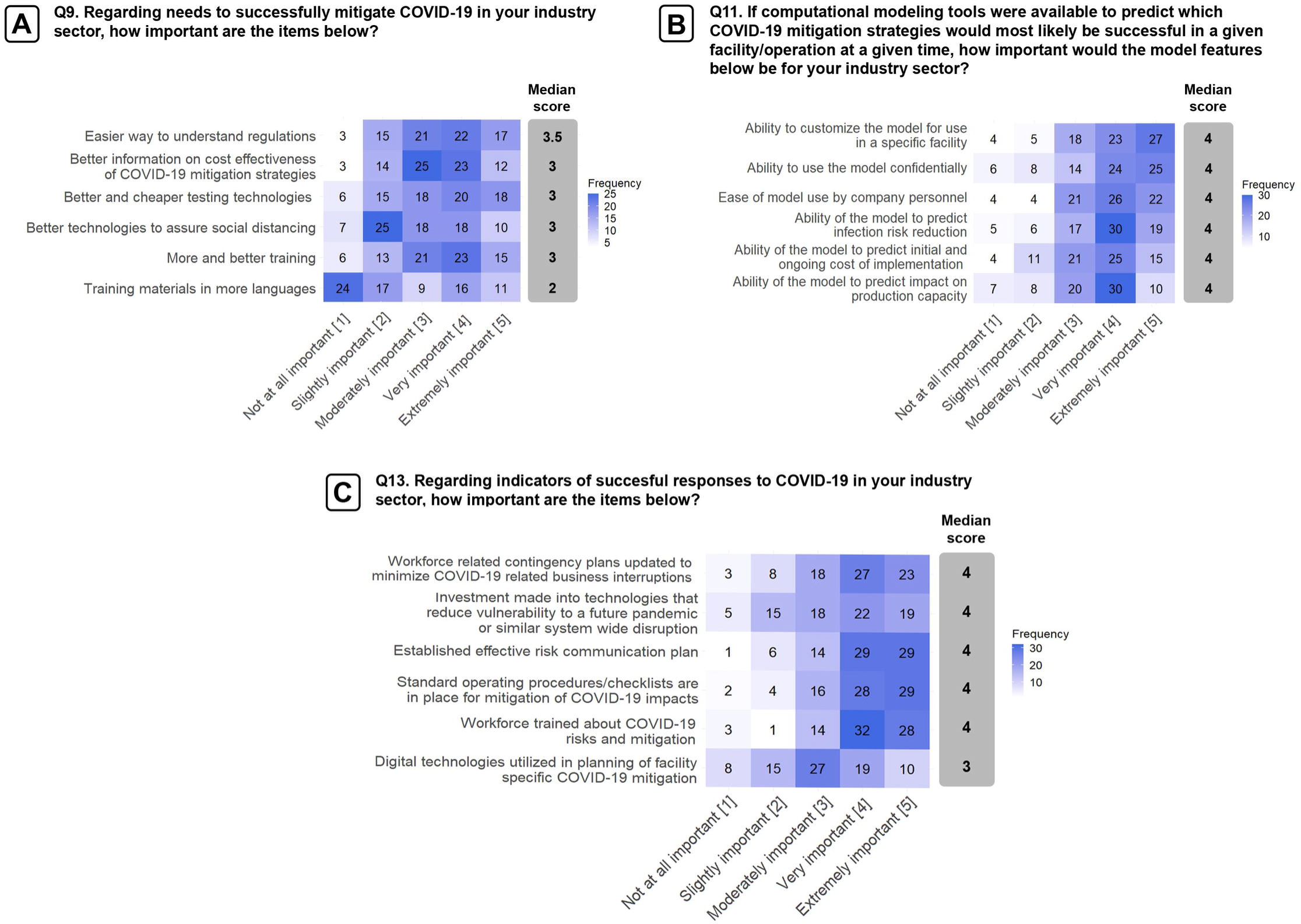
Heatmaps representing responses to 5-point scale Likert questions regarding the [A] needs to successfully mitigate COVID-19, [B] preferred features in predictive models, and [C] indicators of a successful response against COVID-19. These questions were included in part 1 of the needs assessment survey asking general questions about a participant’s industry sector. For each item within a Likert question, a median shown to the right of the heatmap was calculated from the interval values (1-5), assigned to answers ranging from “Not at all important” to “Extremely important”.

If a computational modeling tool were to be available to predict which COVID-19 mitigation strategies would be the most successful in a given facility (Q11, Fig 2B), all model features proposed in the survey (e.g., ease of use, confidentiality and customization, etc.) would be considered very important (median score = 4). Regarding the question about indicators of successful responses to COVID-19 in their industry sector (Q13, Fig 2C), participants perceived that almost all items listed in the question were very important (median score = 4), including establishment of contingency and risk communication plans, standard operating procedures, training, and investment into technologies that reduce vulnerability to a future pandemic (such as due to a new COVID-19 variant) or similar system wide disruption. Additionally, large-sized facilities and operations were more likely to have *Established effective risk communication plans* compared to smaller facilities and operations (Q13, *p* = 0.03, Table 5).

In terms of the risk of shutdown associated with the facility/operation due to work absences for certain specialized job functions (Q27, Fig 3A), participants indicated that *Specialized production line functions*, *Engineering and/or maintenance crew*, and *Sanitation and cleaning* presented a high risk of shutdown (median score = 4). Regarding potential sources of COVID-19 infection in the facility/operation (Q29, Fig 3B), participants were particularly concerned about *Activities in the local community* (median score = 4). In addition, respondents from medium-sized facilities/operations were more likely to be concerned about *Indoor common areas* as a potential source of COVID-19 compared to small and large-sized facilities/operations (Q29, *p* = 0.04 and *p* = 0.05, respectively; Table 5).

**Fig 3.**
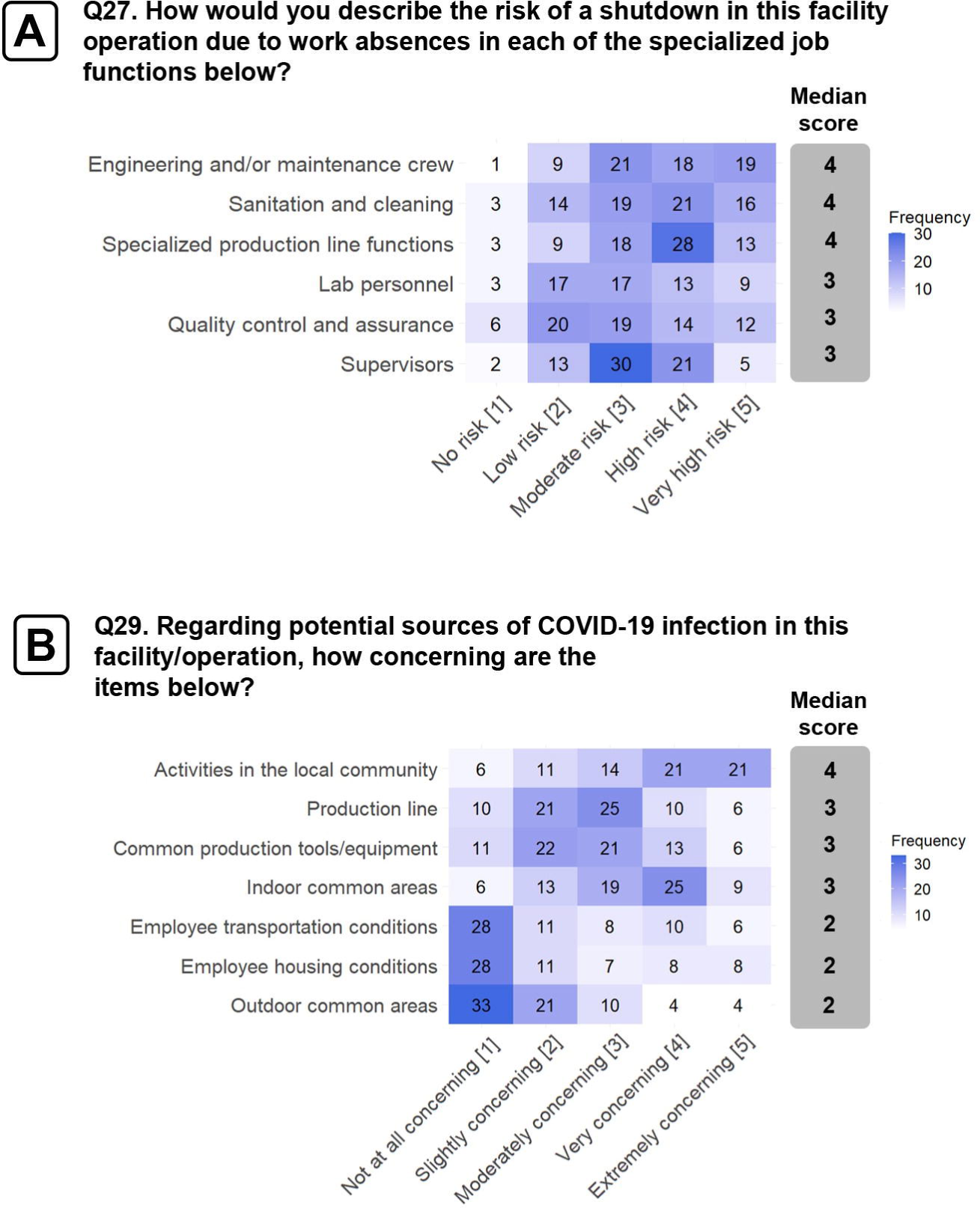
Heatmaps representing responses to 5-point scale Likert questions regarding [A] the risk of shut down due to absenteeism of employees with specialized job functions and [B] sources of COVID-19 in the work environment. These Likert questions were included in part 1 of the needs assessment survey that asked about conditions and COVID-19 controls in a facility or operation of a participant’s choice. For each item within a Likert question, a median shown to the right of the heatmap was calculated from the interval values (1-5), assigned to answers ranging from “No risk” to “Very high risk” [A] and “Not at all concerning” to “Extremely concerning”[B].

### Survey participants self-reported a widespread implementation of social distancing, biosafety, and surveillance mitigation strategies in their facilities/operations representing different sectors of the food industry

Among social distancing measures (Q34, Fig 4A), *Installed Physical barriers*, *Staggered break times*, *Adjusted sick day policy*, and *Spacing workers >6 ft during production* were all implemented or partially/temporarily implemented by most facilities/operations, followed by *Staggered shifts* which were adopted or partially/temporarily adopted by about half of the participants’ facilities/operations. *Downsizing operations* and *Cohorting employees* were found to be the least implemented social distancing strategies to mitigate COVID-19 transmission (Fig 4A). Small facilities/operations implemented several social distancing strategies significantly less frequently than medium- and large-sized facilities and operations, including installing of physical barriers (*p* = 0.005 and *p* = 0.005, respectively), staggered break times (*p* < 0.001 and *p* = 0.004, respectively), and staggered shifts (*p* = 0.02 and *p* = 0.04, respectively;Q34, Table 5). In addition, medium-sized facilities and operations were more likely to adopt *Adjusted sick day policy* (*p* = 0.003) and *Cohorting employees* (*p* = 0.02) as one of their social distancing strategies compared to small-sized facilities and operations (Q34, Table 5). In addition, dairy facilities were more likely to adopt *Spacing workers >6ft during production* (*p* = 0.05) compared to facilities/operations from the group of “Other” industry sectors (Q34, Table 5). The responses to the corresponding open-ended question revealed that infrastructure, productivity, and union- imposed constraints as well as the perceived lack of need or concern were reasons for non-implementation of social distancing measures (thematic analysis of Q35, Table 6). For example, in the following quote one of the participants expressed a concern about downsizing the operation as a strategy to contain the infection spread: “If the plant does not run near full production, we are out of business.” Participant ID #56, Dairy.

**Fig 4.**
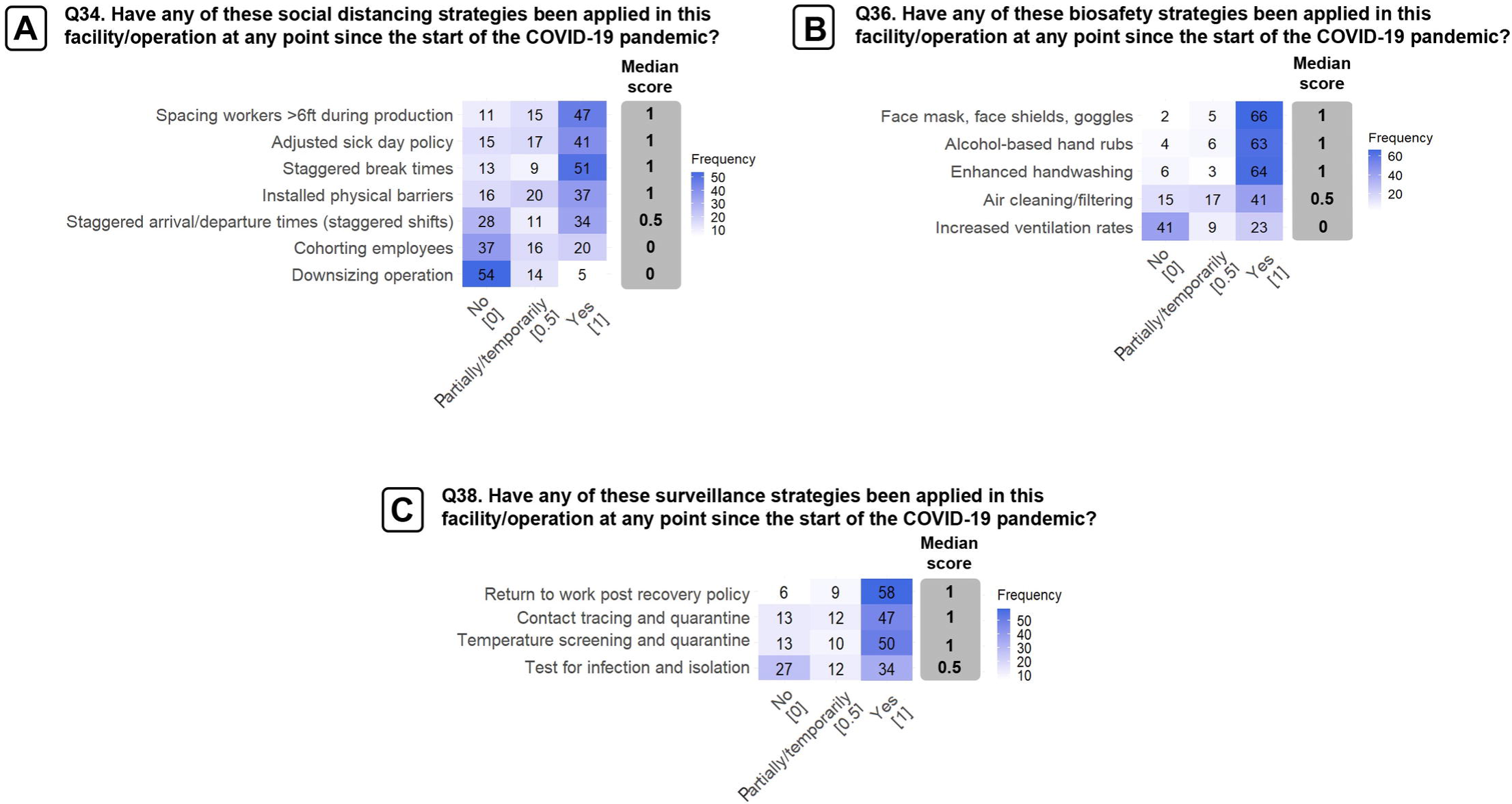
Responses to questions about the implementation of social distancing [A], biosafety [B], and surveillance strategies [C] in a facility/operation of participant’s choice. For each specific mitigation strategy, a median shown to the right of the heatmap was calculated from the numerical values, 0, 0.5 and 1, assigned to answers ‘No’, ‘Yes, but only partially’, and ‘Yes’, respectively.

*Enhanced handwashing*, *Alcohol-based hand rubs*, and *Face mask, face shields, goggles*, were the most implemented biosafety strategies among facilities/operations (Q36, Fig 4B). The latter strategy was implemented more in facilities/operations in the dairy and “Other” industry sector group compared with the fresh produce sector (Q36, *p* = 0.002 and *p* = 0.05, respectively; Table 5). *Air cleaning/filtering* was implemented or partially/temporarily implemented in a little more than half of the facilities/operations, with medium-sized facilities and operations being more likely to adopt it compared to small facilities and operations (Q36, *p* = 0.01; Table 5).

Meanwhile, less than half of the facilities and operations adopted *Increased ventilation rates* as a biosafety measure to mitigate COVID-19 transmission (Q36, Fig 4B). The responses to the corresponding open-ended question indicated that where biosafety interventions were not implemented, it was due to infrastructure constrains or lack of funds, lack of supplies or information, or due to the perceived lack of need (thematic analysis of Q37, Table 6).

Among surveillance strategies (Q38, Fig 4C), *Temperature screening and quarantine*, *Return to work post recovery policy,* and *Contact tracing* were commonly adopted among facilities and operations, while *Test for infection and isolation* was less frequently implemented than the rest of the surveillance strategies based on the comparison of medians of Likert responses. *Contact tracing and quarantine* (*p* < 0.001), *Temperature screening and quarantine* (*p* = 0.006), and *Return to work post recovery policy* (*p* = 0.002) were more commonly adopted in medium-sized compared to small facilities/operations (Q38, Table 5). *Contact tracing and quarantine* were also more commonly implemented in large-sized compared to small facilities/operations (*p <* 0.001). Additionally, dairy facilities more commonly adopted *Contact tracing and quarantine* as a surveillance measure compared to the fresh produce sector (Q38, *p* = 0.02; Table 5). The responses to the corresponding open-ended question stated the cost of implementation and the caused increase in worker absences, as well as the perceived lack of concern or need, as the reasons for no implementation of surveillance strategies (thematic analysis of Q39, Table 6). For example, two participants expressed challenges due to increased absences when implementing temperature screening as a surveillance strategy:

“Temperature screenings aren’t sufficient, as they aren’t always going to detect a mild case. Temperature gun is 30 to 60 dollars. On site Covid tests are borderline unavailable to small and medium sized companies.” Participant ID #56, Dairy.

“We do daily employee temperature readings but have had to isolate many employees for symptoms other than low grade fevers such as backaches, coughing, fatigue, etc.” Participant ID #46, Fresh produce.

## Discussion

This study aimed to identify the needs of the US food industry to maintain production and assure workers’ health during the COVID-19 pandemic and to determine what mitigation strategies are being implemented. Information on this matter is crucial to determine both how the US food industry has already responded to the pandemic and what is still required to enhance its resilience in the face of current and future similar disaster events. The main findings from this study suggest that for the represented sectors of the US food industry: (i) mitigation strategies have been widely implemented in facilities and operations of those who answered our survey, except strategies that reduce productivity, involve major costs, or/and had insufficient information on cost-effectiveness; (ii) facility/operation size and industry sector may impact decision-making regarding implementation of COVID-19 mitigation strategies; and (iii) there are remaining challenges and opportunities to reduce future impacts of COVID-19 and similar disasters. These findings will be discussed in the following paragraphs.

Before discussing these findings, it is important to consider that survey responses were collected from January 2021 to April 2021, a period in which COVID-19 cases in the US food industry were still occurring but there was a declining trend [18]. Additionally, during this time vaccines were becoming available in the US and access was prioritized for essential non-healthcare workers [29], including food industry employees, and the percentage of fully vaccinated people among those eligible ranged from 0 to 21% during that period [30]. Thus, results presented in this study reflect the state of the US food industry sectors in terms of needs and regarding the impact of COVID-19 pandemic on the workforce, food production, and regarding implementation of COVID-19 mitigation in early 2021.

### The US produce growers and food processors widely implemented mitigation strategies except those that would have had negative effects on the production capacity, those that required major investments, and/or those with insufficient information on cost- effectiveness

The study participants self-reported adopting most social distancing mitigation strategies included in the survey, while those measures that negatively impacted production capacity were scarcely adopted. The widespread adoption of most social distancing strategies across assessed food industry sectors is not surprising given such strategies’ well-established key role in reducing the occurrence of COVID-19 cases in different work settings [31] and countries [32]. The continuous promotion of such measures since the beginning of the COVID-19 pandemic, as well as the availability of updated guidance on how to implement them, likely facilitated their adoption by food facilities and operations [22, 33]. For example, a COVID-19 outbreak that occurred in a Colorado mushroom farm on May 6, 2020, was rapidly controlled and further viral dissemination was prevented based on available public health guidance [34]. Additionally, we hypothesize that the common adoption of social distancing mitigation strategies may be partly due to the availability of funding programs launched by the US federal government to financially assist businesses during the pandemic. These programs include the Coronavirus Aid, Relief, and Economic Security Act (CARES Act), the COVID-19 Economic Injury Disaster Loan, and the Coronavirus Food Assistance Program (CFAP), which provided food facilities and operations with financial resources to implement social distancing [35–38]. Despite this, survey participants expressed concerns about financial challenges that limited their ability to install physical barriers, suggesting that additional sources of funding are necessary to further incentivize the adoption of such measures and that companies need to budget in advance for emergency preparedness and responses. Overall, findings from this study suggest that the common adoption of social distancing strategies and the current availability of detailed guidance for their implementation will contribute to rapid implementation of social distancing controls in response to new SARS-CoV-2 beta-coronavirus variants in the current pandemic, future COVID-19 pandemics, and similar disasters. Additionally, our findings address the gap in terms of currently limited information available [15] about the adoption of mitigation strategies in the US food industry. However, it is important to note that representation for the beef/pork and poultry industries was barely achieved in our study, thus future studies on COVID-19 adoption of social distancing mitigation strategies in the US poultry and meat industry sectors are required to complement and update the information reported by Waltenburg [15].

At the same time, our survey revealed that social distancing measures based on cohorting employees and in particular, downsizing operations, were rarely implemented. Intuitively, this may be because businesses avoid strategies that would reduce production capacity or present organizational disruption [39]. Despite findings in this study, downsizing was an option for some businesses during the COVID-19 pandemic to prevent further dissemination of the virus in cases of facility/operation’ outbreaks, even in late 2020 [11]. For instance, capacity reduction in the Canadian beef/pork industry led to financial setbacks to the business and in turn increased the cost to maintain animals for an extended period before slaughter [11]. It has been reported that downsizing operations as well as the increase in sickness-related absences caused an increase in work-related physical demands and job insecurity, which had negative impacts on employees’ mental health [40]. This is relevant to consider, given that one survey participant in this study expressed food industry employees’ struggles with mental health issues after downsizing, particularly due to facilities and operations cutting back management and office staff while distributing the same weight of responsibilities to a smaller workforce. Thus, if facilities/operations plan on adopting, or are forced to adopt, downsizing as a strategy to reduce COVID-19 transmission, measures should be taken not only to prevent downstream food supply disturbances and production losses to the facility but also to protect employees’ mental wellbeing. Further studies are needed to determine the specific economic and other effects of downsizing on the US food industry facilities/operations and their workers prior to and during the COVID-19 pandemic.

Cheap and easy to implement biosafety mitigation strategies were widely adopted by the survey participants’ produce farm operations and food processing facilities, but more information is needed about cost-effectiveness for air cleaning and air filtering/ventilation. The self-reported use of personal protective equipment (PPE) observed in our study agrees with an earlier report from Waltenburg [15], in which 86 out of 111 (77%) of the meat and poultry processing facilities required workers to wear masks. However, this is in contrast with findings by Yung et al. [25], who indicated that between June and July 2020, only a little more than half (20/37, 54%) of consulted dairy farmers in Wisconsin and Minnesota believed that face masks are being used by workers after the COVID-19 pandemic had started. The authors explain that this moderate use of PPE was due to a perceived safeness against COVID-19 transmission in well-ventilated spaces, and because of the difficulties of using such protection in the hot and humid environment of a dairy farm [25]. The discrepancy in findings between Yung et al. [25] and our study could be because the latter assessed dairy processing facilities rather than dairy farms, and targeting the dairy industry in the US instead of being restricted to Wisconsin and Minnesota. Furthermore, survey participants in the current study mentioned that the lack of access to preventive measures, including PPE, was an important concern in efforts to control COVID-19 in their industry sector. Previous reports indicate that the unavailability of supplies early in the COVID-19 pandemic was a severe challenge for businesses, even for front-line essential workers in healthcare institutions in the US [41] and abroad [41, 42]. This shortage in PPE and other hygiene supplies was a consequence of disruptions in the PPE supply chain in the US [43] and elsewhere, whose weaknesses were exposed during this pandemic [44]. The survey findings about the lack of access to preventative measures strongly suggest that establishing a reliable and efficient system to ensure PPE availability and distribution to food facilities/operations is essential for the success of disaster preparedness plans against future pandemics. Altogether, our findings suggest that sectors of the food industry represented in the survey are prepared to implement biosafety strategies in the event of a new variant of the SARS-CoV-2 beta-coronavirus, future COVID-19 pandemics or similar disasters caused by airborne-transmitted pathogens, although the success in their adoption will strongly depend on the establishment of measures to strengthen the local and global supply chain of PPE and other relevant resources.

Contrary to other biosafety strategies, the current study revealed that air filtering and ventilation were scarcely implemented among facilities/operations (at least as of early 2021) due to the lack of or insufficient information about cost-effectiveness and guidance. These findings are somewhat unexpected considering that ventilation and air filtering have been recommended in late 2020 by the CDC as engineering controls (i.e., measures that do not interfere with employees’ work but prevent their interaction with COVID-19) intended to reduce airborne concentrations of SARS-CoV-2 beta-coronavirus in indoor environments [22]. Moreover, poor air quality and air flow inside meat and poultry processing plants have been associated with an increased risk of workers becoming infected with COVID-19 [24]. Reasons for not implementing air filtering expressed by survey participants include the disruption of the controlled environmental conditions required for production, consideration of air filtering as an unnecessary investment when other mitigation strategies are already in place, and the lack of guidance in how to properly apply this strategy. The CDC recognizes the challenges associated with the application of ventilation as a COVID-19 control strategy [22]. Installing air filtering and ventilation systems is indeed a more complex process compared to other recommended biosafety strategies to combat COVID-19 since it requires consideration of several additional factors before their implementation, including the selection of systems adequate to the size, occupancy level, and specific facilities’ features (e.g., production environment) [45]. Because of the complexity of installing air filtering and ventilation systems [22], and the ongoing discussion about the effectiveness of some of these methods in collecting and/or removing viral particles in the air [46], we hypothesize that facility management might favor other, easier to implement and more commonly advised control strategies to prevent COVID-19 transmission. The survey findings highlight the importance of further research and guidance about the implementation of air filtering and ventilation and the advantages in terms of cost-effectiveness they present over other mitigation measures.

Surveillance strategies were widely implemented by the participants’ produce grower operations and food processor facilities/operations with the exception of testing and isolation due to the cost of detecting positive cases of COVID-19. This finding is not surprising considering that both contact tracing and testing, in conjunction with quarantine, have been proposed as useful methods to prevent COVID-19 transmission in the community [47, 48]. These methods have also been promoted by the CDC and OSHA as ways to reduce COVID-19 dissemination in the workplace [33, 49]. Indeed, an early report of mitigation strategies used in the food industry pointed out temperature screening as a widely applied method in the US poultry and meat industry sectors [15], and it is currently being recommended by the CDC to be implemented in the food industry facilities and operations [20]. Nonetheless, some survey participants mentioned difficulties in implementing surveillance mitigation strategies due to funding limitations; this is expected as a significant investment is required to continuously test suspected cases among employees and temporarily remove positives from the workforce. Our findings suggest that the US food industry is willing to apply surveillance measures to prevent further transmission of COVID-19 cases in their facilities/operations and underline the need to improve access to these measures and to strategically apply them to avoid increasing the costs associated with absenteeism and productivity loss.

Regarding temperature screening, some survey participants expressed distrust to use it as an approach to detect mild cases of COVID-19 infection, a topic that has been a matter of discussion throughout the pandemic [50–53]. Slade and Sinha [51] identified issues associated with temperature screening through non-contact infrared thermometers, namely the risk of close interaction between the employees and testers when measuring temperature, the variability in measurements due to lack of training in its application, and the costs associated with hiring employees to continuously measure temperature among workers. Additionally, previous reports have suggested poor specificity [54, 55] and sensitivity [56] of temperature screening (both infrared thermometers and thermal imaging cameras) when used to detect mild COVID-19 cases, which results in many false positives, thus leading to an unnecessarily increased absenteeism and the associated rise in costs due to productivity loss. Chen et al. [55] found that specificity for infrared thermometers can range between 61% to 67%, depending on the part of the body being measured (wrist and forehead, respectively). This is an extremely important drawback for labor-intensive sectors of the food industry, which rely on the availability of qualified workers to continue food production. It was proposed that temperature screening has been a valuable and easy to implement tool to reduce COVID-19 cases, particularly at the beginning of the pandemic when the knowledge about the virus and the effectiveness of preventive methods and available guidance was limited [57, 58]. However, given the issues associated with this strategy, the increased access to vaccination against COVID-19, and the widespread implementation of other controls in the food industry, we believe it relevant to reevaluate the current role and contribution of temperature screening as a surveillance strategy and how it should be implemented (if its implementation is needed at all) to avoid increasing absenteeism among workers. Ideally, surveillance methods should be adopted based on tests’ availability, ease of implementation, and accuracy. For the application of temperature screening, maximizing specificity would be crucial to avoid the costs associated with false-positive cases. This can be achieved by raising the cut-off value used to define a positive case (increasing the temperature cut-off), establishing more stringent criteria for diagnosis of COVID-19 cases (e.g., presence of COVID-19 symptoms, recent exposure to the virus through interaction with infected individuals, or the requirement of follow-up testing for confirmation of suspected individuals), or applying temperature screening in series with a second highly specific diagnostic test.

### Large and medium-sized food industry facilities/operations have better ability to implement mitigation strategies compared to small businesses. Additionally, there are differences among the dairy and fresh produce industry sectors in their needs and adoption of mitigation strategies

Large and medium-sized facilities/operations of survey participants more frequently implemented social distancing, biosafety, and surveillance mitigation strategies compared to small businesses. It is also important to consider that, due to scale economies, implementing mitigation strategies might lead to a greater reduction in production capacity for smaller businesses than for larger ones, resulting in greater economic losses that cannot be sustained by those small businesses, as evidenced by studies pointing out their lack of financial resilience to sustain the impacts of COVID-19 [59, 60]. For example, the implementation of complex measures, such as air cleaning/filtering, which entail high fixed costs, is likely to be economically infeasible or even unnecessary for operations that consist of only a few employees. Findings from the survey reveal that small facilities/operations are more commonly concerned about funding, while participants also mentioned that small facilities/operations struggle to manage COVID-19 related work absences and that better financial support is required to assist small to medium-sized businesses. Because small businesses typically lack access to the types of financial resources readily available to large firms (i.e., institutional funding), research has been directed to develop cutting-edge and low-cost technologies to assist them in responding to the COVID-19 pandemic [61, 62]. Our findings suggest that small food industry facilities and operations would benefit from development of low-cost, effective, and flexible mitigation strategies to help maintain production in their operations during the COVID-19 pandemic.

We found that participants’ facilities from the dairy industry sector more commonly adopted specific biosafety and surveillance mitigation strategies compared to the fresh produce industry sector, while the latter was more likely to be concerned about funding and required easier approaches to understand continuously changing governmental regulations. We hypothesize that the comparatively lower adoption of PPE in the fresh produce industry is due to some operations (farms) being conducted outdoors in the field, in which case the use of goggles and face shields might prevent employees from performing their tasks efficiently. Indeed, in contrast to dairy processing plants, the decentralized nature of the fresh produce industry coupled with the significant heterogeneity in operating practices suggest that not all mitigation strategies may be appropriate for some operations. Additionally, reduced implementation may be due to the fact that transmission risk of COVID-19 outdoors is considered lower than indoors.

Differences may also exist in how consistently both industry sectors can access mitigation strategies, as survey participants from the fresh produce sector expressed concerns about funding limitations. Moreover, our survey findings indicate that the fresh produce industry sector also needed easier approaches to understand the continuously evolving governmental regulations, potentially due to the continuous turnover of workers that are incorporated into the workforce seasonally. These findings reveal important differences in terms of challenges and needs of the dairy and fresh produce industries and further suggest that innovative approaches are needed for fresh produce operations to overcome the limitations in PPE use, as well as develop novel strategies to train the newly recruited workforce more efficiently.

### To reduce the US food industry’s vulnerability, it is important to establish plans and guidelines to minimize business interruptions, train the workforce about risk and mitigation strategies, and develop new technologies to face COVID-19-related disruptions

The survey participants considered the establishment of plans and guidelines as very relevant for reducing the impact of current and future COVID-19 outbreaks. These approaches would enhance industries’ ability to effectively respond to current and future disruptions in a timely manner, considering the consequences of the COVID-19 pandemic in the US food industry, particularly at the beginning of the pandemic [6]. Our findings are consistent with a previous study calling for the food industry to proactively establish plans and mitigation strategies to assist in building resiliency and ensuring the correct continuation of the productive process [63]. This supports the idea that the development of plans and guidelines to prevent future impacts of the COVID-19 pandemic and similar disasters should be established taking into consideration the complexity of the food supply chain. More specifically, government and businesses should develop contingency plans to be prepared to effectively address workforce reduction, economic losses, and facility shutdowns as consequences of the simultaneous effects of dramatic changes in products’ demand [64, 65], modifications to products’ specifications (e.g., package size) [64, 65], government-mandated suspension of operations [64, 65], and facility shutdown-related increase in work absences [6].

The survey participants also acknowledged the importance of training employees in risk and mitigation strategies to prevent COVID-19 transmission in their food facilities and operations. This finding was expected given that training has been a fundamental tool to educate the workforce on how to behave during the pandemic, correctly implement mitigation strategies, and understand the current state and federal rules and regulations around the COVID-19 pandemic. Guidelines and training resources made available by the CDC and OSHA have been useful to train employees in the use of PPE, social distancing practices in the workplace, and identification of signs and symptoms associated with COVID-19 cases. Importantly, ensuring access to the internet in facilities and operations located in remote areas is also crucial to allow them to implement training strategies promptly, a need expressed by survey participants. This is essential if novel technologies, such as augmented and virtual reality, are to be implemented for training the workforce on disaster preparedness [66]. Previous reports evidence important advantages of these novel approaches compared to traditional methods for training, including the opportunity to socially distance while allowing workers to immerse themselves in the activity at hand and learn effectively [67].

The development of new technological approaches to respond to the COVID-19 and future pandemic events (e.g., due to new SARS-CoV-2 beta-coronavirus variants) was considered important by the survey participants. The COVID-19 pandemic has resulted in new technologies being developed at an accelerated pace to promptly assist the food industry in dealing with the COVID-19 pandemic and beyond [66]. Future advances in digitalization and big data analysis will assist the food industry in rapidly responding to COVID-19-related or other similar disruptions through improved access to knowledge for decision-making [5]. For instance, the design of modeling tools to predict impacts of COVID-19, including costs associated with absenteeism, effects on productivity, and the cost-effectiveness of implementing available mitigation strategies would provide useful information to assist the food industry in better managing the ongoing pandemic. Furthermore, improvements in data collection and management, such as shared access to data and data traceability, can facilitate relevant improvements in the efficiency of the food supply chain, while the implementation of automation and robotics technologies could assist in improving food security and facilitate the adoption of social distancing measures to prevent airborne pathogen transmission [5, 65]. Unquestionably, the development of new technologies will assist in building resilience to withstand COVID-19- related disruptions in the US food industry. Nonetheless, we suggest that the implementation of novel technological approaches must consider maintaining previous efforts made by the industry sector to improve efficiency, as well as consider the financial investment necessary to implement technological advances, such as automation [6, 68]. Further studies are needed to determine the specific technological requirements across industry sectors in the US, the ability of food industry sectors to invest in new technologies, and the mechanisms in which these new advances could contribute to improving resilience in the industry.

### Study limitations

The major limitation of the conducted needs assessment survey is relatively low number of responses received, and the associated potential selection bias, despite the engagement of a number of professional and trade organizations and the distribution of the survey via social media. The concurrent widespread application of survey-based studies to understand different aspects of COVID-19 in the US food industry might have resulted in the industry’s fatigue and increased reluctance to respond to surveys as the pandemic progresses, particularly given that the survey designed for this study was distributed more than a year after the pandemic has begun.

This is relevant, as it may have introduced a non-response selection bias to findings presented here, particularly due to the slim response rate from representatives of the beef/pork and no responses from the poultry sectors. Thus, any generalization of findings to these sectors should be done with caution. The reasons for differential participation of the food industry sectors in this study are unknown. Among responses, almost half were from the dairy industry sector, with a much lower representation from the fresh produce industry sector. Similarly, in line with the targeted recruitment efforts, responses were predominantly from individuals in high managerial positions (c-suite, facility managers, etc.), with only a few responses from non-managerial positions. Although this potentially limits the range of perspectives included in the study, opinions, and perceptions from individuals ‘at the top of the ladder’ were targeted because they can provide valuable insight into facilities and operation’s needs, challenges, decisions, and overall impacts of the COVID-19 pandemic in the US food industry. However, we recognize that this study does not include the complete spectrum of worker positions (particularly non-managerial) and therefore not fully address food industry worker needs. Due to the relatively small number of responses, more elaborate approaches to control for potential confounding of the associations between the outcomes and predictors of interest could not be conducted.

Considering Likert-item responses as interval/numerical data during statistical analysis led to the application of non-parametric methods for statistical assessment of associations, which are less efficient for detecting existing effects than parametric tests. The survey was open for participation for almost 3 months in early 2021, which was a time characterized by great changes concerning the response to the COVID-19 pandemic in the US, such as the implementation of COVID-19 vaccination among essential non-healthcare workers. It is possible that some of those changes may have affected participants’ responses depending on when they completed the survey and might not represent needs of the US food industry and adoption of mitigation strategies early in the COVID-19 pandemic. Additionally, although in open-ended questions participants commented about concerns regarding limited access to COVID-19 vaccines for food industry employees and their vaccination hesitancy, the survey did not include questions about vaccination because vaccines were not available at the time the pilot process was finished and the survey was initially distributed. Thus, timely assessment of the food industry needs regarding COVID-19 vaccination is still needed. Finally, although information about COVID-19 morbidity and mortality in the participants’ facilities and operations could have provided valuable insights, we chose to exclude those to improve the response rate.

### Conclusions

Provided that the responses of survey participants are reflective of the wider US food industry, the food industry facilities and operations in the US are broadly prepared to protect their workers and businesses quickly and effectively in the event of a future pandemic due to SARS-CoV-2 or similar airborne pathogens. Future collaborations between the US food industry and federal and state agencies to establish contingency plans and define appropriate training, as well as the development of food industry-directed technologies will be crucial to build resilience against future COVID-19-related and similar disturbances.

## Data Availability

All data (and associated metadata) files analyzed in this study are available for open access in Zenodo via a permanent Digital Object Identifier (DOI: 10.5281/zenodo.5165334)

https://doi.org/10.5281/zenodo.5165290

## Acknowledgments

We thank Alina N. Stelick and Cecil Barnett-Neefs for providing feedback during the needs assessment survey design process. We are also grateful to advisory council members who assisted during the pilot of the needs assessment survey, the professional and trade organizations who assisted in its dissemination, and everyone whose participation made this study possible.

---

S1 Table. All questions (Q) included in the needs assessment survey, question type, type of analysis (statistical or thematic) and role in statistical analysis of associations (as an outcome of interest (primary or secondary) or independent variable (predictor). Responses considered usable for analysis were those in which participants completed at least part 1 of the needs assessment survey.

S2 Table. Themes and underlying codes identified in participants’ responses to selected open-ended questions in the needs assessment survey, which were intended to provide depth to the corresponding Likert questions. An example quote is included to illustrate each of the codes identified.

S3 Table. Significant associations found in the bivariable analyses (P ≤ 0.05) using Mann- Whitney U (MW-U) and Spearman’s correlation (SC) tests between a specific Likert item (outcome) in a survey question (Q) and independent variables (predictors).

S1 Appendix. Copy of the needs assessment survey instrument.

## References

1. Galanakis CM. The food systems in the era of the coronavirus (COVID-19) pandemic crisis. Foods. 2021;9: 523. doi:10.1146/annurev-soc-060116-053252

2. Hayes DJ, Schulz LL, Hart CE, Jacobs KL. A descriptive analysis of the COVID-19 impacts on U.S. pork, turkey, and egg markets. Agribusiness. 2021;37: 122–141. doi:10.1002/agr.21674

3. Singh S, Kumar R, Panchal R, Tiwari MK. Impact of COVID-19 on logistics systems and disruptions in food supply chain. Int J Prod Res. 2021;59: 1993–2008. doi:10.1080/00207543.2020.1792000

4. Barman A, Das R, De PK. Impact of COVID-19 in food supply chain: Disruptions and recovery strategy. Curr Res Behav Sci. 2021;2: 100017. doi:10.1016/j.crbeha.2021.100017

5. Hobbs JE. The Covid-19 pandemic and meat supply chains. Meat Sci. 2021; 108459. doi:10.1016/j.meatsci.2021.108459

6. Weersink A, von Massow M, Bannon N, Ifft J, Maples J, McEwan K, et al. COVID-19 and the agri-food system in the United States and Canada. Agric Syst. 2021;188: 103039. doi:10.1016/j.agsy.2020.103039

7. Qingbin W, Liu C quan, Zhao Y feng, Kitsos A, Cannella M, Wang S kun, et al. Impacts of the COVID-19 pandemic on the dairy industry: Lessons from China and the United States and policy implications. J Integr Agric. 2020;19: 2903–2915. doi:10.1016/S2095-3119(20)63443-8

8. Muth MK. Effects of COVID-19 Meat and Poultry Plant Closures on the Environment and Food Security. 2021 Dec 1 [cited 7 Jul 2021]. In: SESYNC [Internet]. Available from: https://www.sesync.org/news/wed-2020-12-02-1543/effects-of-covid-19-meat-and-poultry-plant-closures-on-the-environment-and

9. Doering C. Meat processors wrestle with worker shortages as US economy reopens from COVID- 19. 2021 June 9 [cited 7 Jun 2021]. In: FOODDIVE [Internet]. Available from: https://www.fooddive.com/news/meat-processors-wrestle-with-worker-shortages-as-us-economy-reopens-from-co/600941/

10. McEwan K, Marchand L, Shang M, Bucknell D. Potential implications of COVID-19 on the Canadian pork industry. Can J Agric Econ. 2020;68: 201–206. doi:10.1111/cjag.12236

11. Hailu G. COVID 19: Balancing and food processing in Canada. Can J Agric Econ Can d’agroeconomie. 2021;69: 177–187. doi:10.1111/cjag.12286

12. Mussell A, Bilyea T, Hedley D. Agri-Food Supply Chains and Covid-19 Resilience and Vulnerability. Agri-Food Econ Syst. 2020; 1–6.

13. Nakat Z, Bou-Mitri C. COVID-19 and the food industry: Readiness assessment. Food Control. 2021;121: 107661. doi:10.1016/j.foodcont.2020.107661

14. Cybersecurity and Infrastructure Security Agency (CISA). Guidance on the Essential Critical Infrastructure Workforce: Ensuring Community and National Resilience in COVID-19 Response. 2020 Aug 18 [cited 7 June 2021]. In: Cybersecurity and Infrastructure Security Agency [Internet]. Available from: https://www.cisa.gov/publication/guidance-essential-critical-infrastructure-workforce

15. Waltenburg M, Victoroff T, Rose CE, Butterfield M, Jervis RH, Fedak KM, et al. Update: COVID-19 Among Workers in Meat and Poultry Processing Facilities - United States, April-May 2020. Report, Mortal Wkly. 2020;69: 557–561.

16. Shelly M. Maine CDC launches investigation into COVID-19 outbreak at Clinton dairy farm. 2021 Dec 7 [cited 7 Jun 2021]. In: Central Maine [Internet]. Available from: https://www.centralmaine.com/2020/12/07/cdc-launches-investigation-into-covid-19-outbreak-at-clinton-dairy-farm/

17. Kielar M. Lessons learned from one of the largest COVID-19 outbreaks in New York. 2021 Nov 9 [cited 2 Jun 2021]. In: CNYCENTRAL [Internet]. Available from: https://cnycentral.com/news/local/investigation-reveals-lessons-learned-from-one-of-the-largest-covid-19-outbreaks-in-nys

18. Douglas L. Mapping Covid-19 outbreaks in the food system. 2020 Apr 22 [cited 7 Jun 2021]. In: Food and Environment Reporting Network [Internet]. Available from: https://thefern.org/2020/04/mapping-covid-19-in-meat-and-food-processing-plants/

19. Chen YH, Glymour M, Riley A, Balmes J, Duchowny K, Harrison R, et al. Excess mortality associated with the COVID-19 pandemic among Californians 18-65 years of age, by occupational sector and occupation: March through November 2020. PLoS One. 2021;16: 1–10. doi:10.1371/journal.pone.0252454

20. Centers for Disease Control and Prevention (CDC). Agriculture Workers and Employers. 2020 June 2 [cited June 18 2021]. In: Centers for Disease Control and Prevention [Internet]. Available from: https://www.cdc.gov/coronavirus/2019-ncov/community/guidance-agricultural-workers.html

21. Occupational Safety and Health Administration (OSHA). Protecting workers: Guidance on mitigating and preventing the spread of COVID-19 in the workplace. 2021 Jan 29 [cited June 18, 2021]. In: Occupational Safety and Health Administration [Internet]. Available from: https://www.osha.gov/coronavirus/safework

22. Centers for Disease Control and Prevention (CDC). Ventilation in Buildings. 2020 Dec 7 [cited June 18 2021]. In: Centers for Disease Control and Prevention [Internet]. Available from: https://www.cdc.gov/coronavirus/2019-ncov/community/ventilation.html

23. Occupational Safety and Health Administration (OSHA), Food and Drug administration (FDA). Employee Health and Food Safety Checklist for Human and Animal Food Operations During the COVID-19 Pandemic. 2020 Aug 11 [cited June 18 2021]. In: Occupational Safety and Health Administration [Internet]. Available from: https://www.fda.gov/media/141141/download

24. Pokora R, Kutschbach S, Weigl M, Braun D, Epple A, Lorenz E, et al. Investigation of superspreading COVID-19 outbreak events in meat and poultry processing plants in Germany: A cross-sectional study. PLoS One. 2021;16: 1–14. doi:10.1371/journal.pone.0242456

25. Yung MT, Vázquez RI, Liebman A, Brihn A, Olson A, Loken D, et al. COVID-19 Awareness and Preparedness of Minnesota and Wisconsin Dairy Farms. J Agromedicine. 2021;26: 352–359. doi:10.1080/1059924X.2021.1927925

26. R Core Team. R: A language and environment for statistical computing. R Foundation for Statistical Computing. 2017. Available from: https://www.r-project.org/

27. Benjamini J, Hochberg Y. Controlling the False Discovery Rate : A Practical and Powerful Approach to Multiple Testing. J R Stat Soc Ser B. 1995;57: 289–300.

28. Clarke V, Braun V. Thematic Analysis. In: Teo T, editor. Encyclopedia of Critical Psychology. New York, NY: Springer New York; 2014. pp. 1947–1952. doi:10.1007/978-1-4614-5583-7_311

29. Centers for Disease Control and Prevention (CDC). Interim List of Categories of Essential Workers Mapped to Standardized Industry Codes and Titles. 2020 Dec 22 [cited 2 Aug 2021]. In: Centers for Disease Control and Prevention [Internet]. Available from: https://www.cdc.gov/vaccines/covid-19/categories-essential-workers.html

30. John Hopkins University. The vaccine story. 2021 May 16 [cited Jun 7 2021]. In: John Hopkins University [Internet]. Available: https://coronavirus.jhu.edu/vaccines/story

31. Kluger DM, Aizenbud Y, Jaffe A, Parisi F, Aizenbud L, Minsky-Fenick E, et al. Impact of healthcare worker shift scheduling on workforce preservation during the COVID-19 pandemic. Infect Control Hosp Epidemiol. 2020;41: 1443–1445. doi:10.1017/ice.2020.337

32. Thu TPB, Ngoc PNH, Hai NM, Tuan LA. Effect of the social distancing measures on the spread of COVID-19 in 10 highly infected countries. Sci Total Environ. 2020;742: 140430. doi:10.1016/j.scitotenv.2020.140430

33. Occupational Safety and Health Administration (OSHA). Guidance on Returning to Work. 2020 Jun 17 [cited Jun 17 2021]. In: Occupational Safety and Health Administration [Internet]. Available from: https://www.osha.gov/Publications/OSHA4045.pdf

34. Alamosa News. Mushroom farm confirms COVID-19 cases. 2020 May 9 [cited 29 Jun 2020]. In: Alamosa News [Internet]. Available from: https://alamosanews.com/article/mushroom-farm-confirms-covid-19-cases

35. U.S. Department of Agriculture (USDA). USDA Announces Coronavirus Food Assistance Program. 2020 Apr 17 [cited 17 Jun 2021]. In: U.S. Department of Agriculture [Internet]. Available from: https://www.usda.gov/media/press-releases/2020/04/17/usda-announces-coronavirus-food-assistance-program

36. U.S. Small Business Administration (SBA). COVID-19 EIDL. 2021 Jun 16 [cited 17 Jun 2021. Available from: U.S. Small Business Administration [Internet]. https://www.sba.gov/funding-programs/loans/covid-19-relief-options/eidl/covid-19-eidl

37. U.S. Congress. H.R.748 - CARES Act. 2020 March 27 [cited Jun 17 2021]. In: U.S. Congress [Internet]. Available from: https://www.congress.gov/bill/116th-congress/house-bill/748

38. U.S. Small Business Administration (SBA). Paycheck Protection Program. 2020 Mar 20. In: U.S. Small Business Administration [Internet]. Available from: https://www.sba.gov/funding-programs/loans/covid-19-relief-options/paycheck-protection-program

39. Zorn ML, Norman PM, Butler FC, Bhussar MS. Cure or curse: Does downsizing increase the likelihood of bankruptcy? J Bus Res. 2017;76: 24–33. doi:10.1016/j.jbusres.2017.03.006

40. Kivimäki M, Vahtera J, Pentti J, Ferrie JE. Factors underlying the effect of organisational downsizing on health of employees: Longitudinal cohort study. Br Med J. 2000;320: 971– 975. doi:10.1136/bmj.320.7240.971

41. Ahmed J, Malik F, Bin Arif T, Majid Z, Chaudhary MA, Ahmad J, et al. Availability of Personal Protective Equipment (PPE) Among US and Pakistani Doctors in COVID-19 Pandemic. Cureus. 2020;12: e8550. doi:10.7759/cureus.8550

42. Kim H, Hegde S, Lafiura C, Raghavan M, Sun N, Cheng S, et al. Access to personal protective equipment in exposed healthcare workers and COVID-19 illness, severity, symptoms and duration: A population-based case-control study in six countries. BMJ Glob Heal. 2021;6: 1–9. doi:10.1136/bmjgh-2020-004611

43. Cohen J, Rodgers Y van der M. Contributing factors to personal protective equipment shortages during the COVID-19 pandemic. Prev Med (Baltim). 2020;141: 106263. doi:10.1016/j.ypmed.2020.106263

44. Dai T, Zaman MH, Padula W V., Davidson PM. Supply chain failures amid Covid-19 signal a new pillar for global health preparedness. J Clin Nurs. 2021;30: e1–e3. doi:10.1111/jocn.15400

45. Blocken B, van Druenen T, Ricci A, Kang L, van Hooff T, Qin P, et al. Ventilation and air cleaning to limit aerosol particle concentrations in a gym during the COVID-19 pandemic. Build Environ. 2021;193: 107659. doi:10.1016/j.buildenv.2021.107659

46. Ham S. Prevention of exposure to and spread of COVID-19 using air purifiers: challenges and concerns. Epidemiol Health. 2020;42: 1–3. doi:10.4178/epih.e2020027

47. Aleta A, Martín-Corral D, Pastore y Piontti A, Ajelli M, Litvinova M, Chinazzi M, et al. Modelling the impact of testing, contact tracing and household quarantine on second waves of COVID-19. Nat Hum Behav. 2020;4: 964–971. doi:10.1038/s41562-020-0931-9

48. Quilty BJ, Clifford S, Hellewell J, Russell TW, Kucharski AJ, Flasche S, et al. Quarantine and testing strategies in contact tracing for SARS-CoV-2: a modelling study. Lancet Public Heal. 2021;6: e175–e183. doi:10.1016/S2468-2667(20)30308-X

49. Centers for Disease Control and Prevention (CDC). SARS-CoV-2 testing strategy: considerations for non-healthcare workplaces. 2021 Jul 4 [cited July 17 2021]. In: Centers for Disease Control and Prevention [Internet]. Available from: https://www.cdc.gov/coronavirus/2019-ncov/community/organizations/testing-non-healthcare-workplaces.html

50. Bielecki M, Crameri GAG, Schlagenhauf P, Buehrer TW, Deuel JW. Body temperature screening to identify SARS-CoV-2 infected young adult travellers is ineffective. Travel Med Infect Dis. 2020;37: 101832. doi:10.1016/j.tmaid.2020.101832

51. Slade DH, Sinha MS. Return to Work during COVID-19: Temperature Screening is No Panacea. Infect Control Hosp Epidemiol. 2020;2019: 1–2. doi:10.1017/ice.2020.1225

52. Mitra B, Luckhoff C, Mitchell RD, O’Reilly GM, Smit DV, Cameron PA. Temperature screening has negligible value for control of COVID-19. Emerg Med Australas. 2020;32: 867–869. doi:10.1111/1742-6723.13578

53. Facente SN, Hunter LA, Packel LJ, Li Y, Harte A, Nicolette G, et al. Feasibility and effectiveness of daily temperature screening to detect COVID-19 in a large public university setting. medRxiv: [Preprint]. 2021. doi: 10.1101/2021.03.22.21254140

54. Martinez-Jimenez MA, Loza-Gonzalez VM, Kolosovas-Machuca ES, Yanes-Lane ME, Ramirez-GarciaLuna AS, Ramirez-GarciaLuna JL. Diagnostic accuracy of infrared thermal imaging for detecting COVID-19 infection in minimally symptomatic patients. Eur J Clin Invest. 2021;51: 1–8. doi:10.1111/eci.13474

55. Chen G, Xie J, Dai G, Zheng P, Hu X, Lu H, et al. Validity of the use of wrist and forehead temperatures in screening the general population for covid-19: A prospective real-world study. Iran J Public Health. 2020;49: 57–66. doi:10.18502/ijph.v49is1.3670

56. Pană BC, Lopes H, Furtunescu F, Franco D, Rapcea A, Stanca M, et al. Real-World Evidence: The Low Validity of Temperature Screening for COVID-19 Triage. Front Public Heal. 2021;9: 1–6. doi:10.3389/fpubh.2021.672698

57. Daga MK. From SARS-CoV to Coronavirus Disease 2019 (COVID-19) - A Brief Review. J Adv Res Med. 2020;06: 1–9. doi:10.24321/2349.7181.201917

58. Izzetti R, Nisi M, Gabriele M, Graziani F. COVID-19 Transmission in Dental Practice: Brief Review of Preventive Measures in Italy. J Dent Res. 2020;99: 1030–1038. doi:10.1177/0022034520920580

59. Bartik AW, Bertrand M, Cullen Z, Glaeser EL, Luca M, Stanton C. The impact of COVID- 19 on small business outcomes and expectations. Proc Natl Acad Sci U S A. 2020;117: 17656–17666. doi:10.1073/pnas.2006991117

60. Fairlie R. The impact of COVID-19 on small business owners: Evidence from the first three months after widespread social-distancing restrictions. J Econ Manag Strateg. 2020;29: 727–740. doi:10.1111/jems.12400

61. Akpan IJ, Udoh EAP, Adebisi B. Small business awareness and adoption of state-of-the-art technologies in emerging and developing markets, and lessons from the COVID-19 pandemic. J Small Bus Entrep. 2020;0: 1–18. doi:10.1080/08276331.2020.1820185

62. Akpan IJ, Soopramanien D, Kwak DH. Cutting-edge technologies for small business and innovation in the era of COVID-19 global health pandemic. J Small Bus Entrep. 2020;0: 1– 11. doi:10.1080/08276331.2020.1799294

63. Boyac -Gündüz CP, Ibrahim SA, Wei OC, Galanakis CM. Transformation of the food sector: Security and resilience during the covid-19 pandemic. Foods. 2021. doi:10.3390/foods10030497

64. Luckstead J, Nayga RM, Snell HA. Labor Issues in the Food Supply Chain Amid the COVID-19 Pandemic. Appl Econ Perspect Policy. 2021;43: 382–400. doi:10.1002/aepp.13090

65. Hobbs JE. Food supply chain resilience and the COVID-19 pandemic: What have we learned? Can J Agric Econ. 2021; 189–196. doi:10.1111/cjag.12279

66. Galanakis CM, Rizou M, Aldawoud TMS, Ucak I, Rowan NJ. Innovations and technology disruptions in the food sector within the COVID-19 pandemic and post-lockdown era. Trends Food Sci Technol. 2021;110: 193–200. doi:10.1016/j.tifs.2021.02.002

67. Akçayır M, Akçayır G. Advantages and challenges associated with augmented reality for education: A systematic review of the literature. Educ Res Rev. 2017;20: 1–11. doi:10.1016/j.edurev.2016.11.002

68. Saitone TL, Aleks Schaefer K, Scheitrum DP. COVID-19 morbidity and mortality in U.S. meatpacking counties. Food Policy. 2021;101: 102072. doi:10.1016/j.foodpol.2021.102072

