## Supplementary material for "Ongoing mitigation strategies and further needs of the United States food industry to control COVID-19 in the work environment": S1 Appendix

**Enclosed is a questionnaire survey instrument used in this study**

We are asking you to participate in a research study titled *Modeling and training to enhance resilience of the US food system to COVID-19 labor shortages*. We will describe this study to you and answer any of your questions. This study is being led by Dr. Renata Ivanek at Cornell University.

**What the study is about**
The purpose of this research is to assess the most urgent needs of the produce, dairy, beef/pork, and poultry sectors of the food industry with respect to the COVID-19 pandemic to be able to assure workers' health and maintain food production with well-targeted, food sector specific and optimal mitigation plans.

**Eligibility**
Participants must be older than 18 years of age and affiliated with a fresh produce operation or with a produce, dairy, beef/pork, poultry or other food processing facility.

**What we will ask you to do**
We will ask you to answer an online survey regarding your perceptions about COVID-19 impacts and control in the food industry. The survey takes approximately 30 minutes to complete.

**Risks and discomforts**
We anticipate that your participation in this survey presents no greater risk than everyday use of the Internet.

**Benefits**
Your participation in this survey will help us learn more about the needs of the food industry with respect to COVID-19, to be able to identify optimal COVID-19 control strategies and develop effective training materials for the food industry.

**Compensation for participation**
There will be no compensation for participation.

**Privacy/Confidentiality/Data Security**
No identifying data will be collected meaning that the survey will be completely anonymous.

**Sharing Anonymous Data Collected in this Research**
Anonymous (non-identifying) data from this study may be shared with the research community at large to advance science and health. As no personal information will be collected, by current scientific standards and known methods, no one will be able to identify you from the information we share. Despite these measures, we cannot guarantee your anonymity.

**Taking part is voluntary**
Participation in this survey is voluntary. If you decide not to participate in this study, it will not affect your current or future relationship with Cornell University. By completing this survey, you are voluntarily agreeing to participate. You may choose to answer all or only a portion of the survey, however, we encourage you to answer as many questions as possible so as to increase the value of the information obtained from this study. Several questions in the survey, that are considered essential for this study, require answers to be able to complete the survey. Questions for which this applies are clearly indicated in the survey. You are free to choose not to participate if you are uncomfortable with this condition. If you decide to participate in this study, please complete the electronic questionnaire you will be able to access after providing your consent below.

**If you have questions**
The main researcher conducting this study is Dr. Renata Ivanek, a professor at Cornell University. For any questions you have now or later, please contact Dr. Renata Ivanek at . If you have any questions or concerns regarding your rights as a subject in this study, you may contact the Institutional Review Board (IRB) for Human Participants at 607-255-5138 or access their website at http://www.irb.cornell.edu. You may also report your concerns or complaints anonymously through

Ethicspoint online at www.hotline.cornell.edu or by calling toll free at 1-866-293-3077. Ethicspoint is an independent organization that serves as a liaison between the University and the person bringing the complaint so that anonymity can be ensured.

**Statement of Consent**
To acknowledge that you have read the information provided above and have received answers to any of your questions, and to consent to take part in the study, please check the box below.

**☐ I have read the above description of this study and I am aware about any risks or benefits involved in my participation**

**This needs assessment has two (2) parts:**

- **Part 1** is about **your overall food industry sector**.
- **Part 2** is about **your specific food production facility/operation**.

**PART 1**
**Block 1:** General questions about your **food industry sector**
Q1. What **industry sector** are you in? (select all that apply)
**(Response to this question is required)**

☐ Fresh produce
☐ Dairy
☐ Beef/Pork
☐ Poultry
☐ Other: ________

Q2. Select your **main role** within your organization.
**(Response to this question is required)**

O C-suite
O Regional manager
O Facility manager
O Research and development
O Corporate food safety and quality
O Other: ________
O Prefer not to answer

Q3. Did COVID-19 have a **significant impact** on your industry sector?

O Yes
O No

**[If Q3 = Yes, display Q4]**
Q4. In which way(s) has COVID-19 **significantly impacted** your industry sector? (select all that apply)

☐ Operations/production has been reduced/cut back
☐ Operations/production has expanded
☐ Implemented robotics, sensors, automation, and/or computer modeling
☐ Management/corporate employees working remotely
☐ Major changes in operational staffing control and protection protocols
☐ Other: ________

Q5. Regarding control of COVID-19 in your industry sector, **how concerning** are the items below?

| | Do not know | 1 - Not at all concerning | 2 - Slightly concerning | 3 - Moderately concerning | 4 - Very concerning | 5 - Extremely concerning |
|---|---|---|---|---|---|---|
| Organizational awareness of the virus | ☐ | ☐ | ☐ | ☐ | ☐ | ☐ |
| Labor availability | ☐ | ☐ | ☐ | ☐ | ☐ | ☐ |
| Workers' compliance with control measures | ☐ | ☐ | ☐ | ☐ | ☐ | ☐ |
| Workers' abuse of control measures | ☐ | ☐ | ☐ | ☐ | ☐ | ☐ |
| Limited financial resources | ☐ | ☐ | ☐ | ☐ | ☐ | ☐ |
| Production capacity | ☐ | ☐ | ☐ | ☐ | ☐ | ☐ |
| Product quality | ☐ | ☐ | ☐ | ☐ | ☐ | ☐ |
| Supplier management | ☐ | ☐ | ☐ | ☐ | ☐ | ☐ |
| Customer expectations | ☐ | ☐ | ☐ | ☐ | ☐ | ☐ |
| Complex/ever-changing government regulations about COVID-19 | ☐ | ☐ | ☐ | ☐ | ☐ | ☐ |

Q6. Regarding control of COVID-19 in your industry sector, are there **any other concerns** we should consider?

____________________________________________________________
____________________________________________________________

Q7. Regarding the labor force needed to maintain the production capacity in your industry sector during the COVID-19 pandemic, **how challenging** are the items below?

| | Do not know | 1 - Not at all challenging | 2 - Slightly challenging | 3 - Moderately challenging | 4 - Very challenging | 5 - Extremely challenging |
|---|---|---|---|---|---|---|
| Access to number of workers needed | ☐ | ☐ | ☐ | ☐ | ☐ | ☐ |
| Access to workers with necessary skills | ☐ | ☐ | ☐ | ☐ | ☐ | ☐ |
| Sufficient housing for labor | ☐ | ☐ | ☐ | ☐ | ☐ | ☐ |
| Turnover in workforce | ☐ | ☐ | ☐ | ☐ | ☐ | ☐ |
| Need to train labor | ☐ | ☐ | ☐ | ☐ | ☐ | ☐ |

Q8. Regarding the labor force needed to maintain the production capacity in your industry sector during the COVID-19 pandemic, are there **any other challenges** we should consider?

________________________________________________________________

________________________________________________________________

Q9. Regarding needs to successfully mitigate COVID-19 in your industry sector, **how important** are the items below?
**(Response to this question is required)**

| | Do not know (1) | 1 - Not at all important (6) | 2 - Slightly important (7) | 3 - Moderately important (2) | 4 - Very important (8) | 5 - Extremely important (9) |
|---|---|---|---|---|---|---|
| More and better training | ☐ | ☐ | ☐ | ☐ | ☐ | ☐ |
| Training materials in more languages | ☐ | ☐ | ☐ | ☐ | ☐ | ☐ |
| Better technologies to assure social distancing | ☐ | ☐ | ☐ | ☐ | ☐ | ☐ |
| Better and cheaper testing technologies | ☐ | ☐ | ☐ | ☐ | ☐ | ☐ |
| Easier way to understand regulations | ☐ | ☐ | ☐ | ☐ | ☐ | ☐ |
| Better information on cost effectiveness of COVID-19 mitigation strategies | ☐ | ☐ | ☐ | ☐ | ☐ | ☐ |

Q10. Regarding needs to successfully mitigate COVID-19 in your industry sector, are there **any other important needs** we should consider?

________________________________________________________________

________________________________________________________________

Q11. If computational modeling tools were available to predict which COVID-19 mitigation strategies would most likely be successful in a given facility/operation at a given time, **how important** would the model features below be for your industry sector?
**Note: this question is about priorities for development of models; answers will not necessarily represent your business priorities.** For example, a ranking of "extremely important" for "Ability of the model to predict impact on production capacity" would not mean that company prioritizes production capacity over worker health; it simply indicates that availability of a model that predicts the impact of COVID-19 interventions on production capacity is a high priority.
**(Response to this question is required)**

| | Do not know | 1 - Not at all important | 2 - Slightly important | 3 - Moderately important | 4 - Very important | 5 - Extremely important |
|---|---|---|---|---|---|---|
| Ability of the model to predict impact on production capacity | ☐ | ☐ | ☐ | ☐ | ☐ | ☐ |
| Ability of the model to predict initial and ongoing cost of implementation | ☐ | ☐ | ☐ | ☐ | ☐ | ☐ |
| Ability of the model to predict infection risk reduction | ☐ | ☐ | ☐ | ☐ | ☐ | ☐ |
| Ease of model use by company personnel | ☐ | ☐ | ☐ | ☐ | ☐ | ☐ |
| Ability to use the model confidentially | ☐ | ☐ | ☐ | ☐ | ☐ | ☐ |
| Ability to customize the model for use in a specific facility | ☐ | ☐ | ☐ | ☐ | ☐ | ☐ |

Q12. Regarding a potential modelling tool that could predict successful COVID-19 mitigation strategies, are there **any other model features important** for your industry sector we should consider?

____________________________________________

____________________________________________

Q13. Regarding indicators of successful responses to COVID-19 in your industry sector, **how important** are the items below?
**(Response to this question is required)**

| | Do not know | 1 - Not at all important | 2 - Slightly important | 3 - Moderately important | 4 - Very important | 5 - Extremely important |
|---|---|---|---|---|---|---|
| Workforce trained about COVID-19 risks and mitigation | ☐ | ☐ | ☐ | ☐ | ☐ | ☐ |
| Standard operating procedures/checklists are in place for mitigation of COVID-19 impacts | ☐ | ☐ | ☐ | ☐ | ☐ | ☐ |
| Established effective risk communication plan | ☐ | ☐ | ☐ | ☐ | ☐ | ☐ |
| Digital technologies utilized in planning of facility specific COVID-19 mitigation | ☐ | ☐ | ☐ | ☐ | ☐ | ☐ |
| Investment made into technologies that reduce vulnerability to a future pandemic or similar system wide disruption | ☐ | ☐ | ☐ | ☐ | ☐ | ☐ |
| Workforce related contingency plans updated to minimize COVID-19 related business interruptions | ☐ | ☐ | ☐ | ☐ | ☐ | ☐ |

Q14. Regarding attainable indicators of successful responses to COVID-19 in your industry sector, are there **any other important indicators** we should consider?

______________________________________________________________________

______________________________________________________________________

**PART 2**

**The remaining questions in this needs assessment ask about conditions and COVID-19 mitigation in your food production facility/operation in the USA**. If you oversee multiple facilities/operations, please provide information about one facility/operation of your choice.

**Block 2:** Questions about **your facility/operation *in 2019***

Q15. In what **industry sector** is this facility/operation?

**(Response to this question is required)**

O Fresh produce

O Dairy

O Beef/Pork

O Poultry

O Other: _________

**[If Q15 = Fresh produce, display Q16]**

Q16. How does this facility/operation operate?

O Year round

O Seasonally

**[If Q16 = Seasonally, display Q17]**

Q17. What are the approximate start and end dates for the **production season(s)** in this fresh produce facility/operation?

Season start and end date(s) ____________________________________________________

**[If Q15 = Fresh produce, display Q18]**

Q18. What **role** best describes this facility/operation?

O Grower

O Packing House

O Processor

O Grower and Field packer

O Grower and Processor

O Other: _________

**[If Q18 = Grower and Processor, display Q19]**

Q19. Please select which **part** of your Grower and Processor operation will you describe in the remaining questions?

O Grower operation

O Processor facility

Q20. What was the **average number of employees** in this facility/operation in 2019?

O Less than 10

O 10-49

O 50-99

O 100-249

O 250-499

O 500-999

O 1000-2000

O More than 2000

O This facility/operation did not operate in 2019

O Prefer not to answer

Q21. What is the approximate proportion (%) of employees in this facility/operation that are **between 50-69 years of age and 70 years old or older**?

Proportion (%) of 50-69 years old ______________________________

Proportion (%) of 70 years old or older ______________________________

Q22. Does this facility/operation provide **group temporary (seasonal) housing** to any of your employees?

O Yes

O No

**[If Q22 = Yes, display Q23]**
Q23. Approximately what proportion (%) of employees in this facility/operation are provided with **group temporary housing**?

Temporary housing provided to this proportion (%) of employees:

________________________________________

Q24. Does this facility/operation **provide group transportation services** (bus, truck, etc.) to employees to/from work?

O Yes

O No

**[If Q24 = Yes, display Q25]**
Q25. Approximately what proportion (%) of employees in this facility/operation are provided with **group transportation** to/from work?

Transportation provided to this proportion (%) of employees

________________________________________

**<u>Block 3</u>:** Questions about conditions in **your facility/operation *in 2020***
Q26. What is the **largest percent reduction in the general production labor force** that this facility/operation could withstand over a period of one week without reduction in the production capacity? **(Response to this question is required)**

O 5%

O 10%

O 15%

O 20%

O 30%

O 40%

O 50%

O Do not know

Q27. COVID-19 related work absences among workers performing different specialized job functions in this facility/operation present different levels of risk for a facility/operation shutdown. How would you describe the **risk of a shutdown** in this facility/operation due to work absences in each of the specialized job functions below?

| | Not applicable | 1 - No risk | 2 - Low risk | 3 - Moderate risk | 4 - High risk | 5 - Very high risk |
|---|---|---|---|---|---|---|
| Specialized production line functions (for example, operators for specialized equipment) | ☐ | ☐ | ☐ | ☐ | ☐ | ☐ |
| Lab personnel | ☐ | ☐ | ☐ | ☐ | ☐ | ☐ |
| Quality control and assurance | ☐ | ☐ | ☐ | ☐ | ☐ | ☐ |
| Sanitation and cleaning | ☐ | ☐ | ☐ | ☐ | ☐ | ☐ |
| Supervisors | ☐ | ☐ | ☐ | ☐ | ☐ | ☐ |
| Engineering and/or maintenance crew | ☐ | ☐ | ☐ | ☐ | ☐ | ☐ |

Q28. Regarding the risk of a shutdown in this facility/operation due to work absences, are there **any other specialized job functions** we should consider?

______________________________________________________________________

______________________________________________________________________

Q29. Regarding potential sources of COVID-19 infection in this facility/operation, **how concerning** are the items below?

| | Do not know | 1 - Not at all concerning | 2 - Slightly concerning | 3 - Moderately concerning | 4 - Very concerning | 5 - Extremely concerning |
|---|---|---|---|---|---|---|
| Employee housing conditions | ☐ | ☐ | ☐ | ☐ | ☐ | ☐ |
| Employee transportation conditions | ☐ | ☐ | ☐ | ☐ | ☐ | ☐ |
| Indoor common areas (for example, restrooms, breakrooms, personal protective equipment area, offices) | ☐ | ☐ | ☐ | ☐ | ☐ | ☐ |
| Outdoor common areas | ☐ | ☐ | ☐ | ☐ | ☐ | ☐ |
| Common production tools/equipment | ☐ | ☐ | ☐ | ☐ | ☐ | ☐ |
| Production line | ☐ | ☐ | ☐ | ☐ | ☐ | ☐ |
| Activities in the local community | ☐ | ☐ | ☐ | ☐ | ☐ | ☐ |

Q30. Regarding potential sources of COVID-19 infection in this facility/operation, are there **any other concerns** we should consider?

______________________________________________________________________

______________________________________________________________________

Q31. If in the future the available labor force in this facility/operation is affected by a COVID-19 outbreak, what **short/mid-term solutions** should be considered to maintain production? (check all that apply)

☐ Extend the number of work hours for remaining workers

☐ Backfill with emergency personnel from third-party companies

☐ Backfill by reorganizing personnel in the same facility

Q32. To prevent labor shortage in this facility/operation due to a potential future COVID-19 outbreak, should **capital investment into mechanization** be considered as a long-term solution to maintain production?

O Yes

O No

Q33. To maintain production in the event of labor shortage in this facility/operation due to a potential future COVID-19 outbreak, are there **any other solutions** we should consider?

______________________________________________________________________________

______________________________________________________________________________

**<u>Block 4</u>:** Questions about different **social distancing** strategies for mitigation of COVID-19 in your facility/operation

***Definitions of social distancing mitigation strategies:***

- **Installed physical barriers:** Clear plastic partitions preventing employees from getting too close and preventing particles or droplets exhaled by one person from entering the breathing zone of another.
- **Staggered break times:** Groups of employees have different break times.
- **Staggered arrival/departure times (staggered shifts):** Groups of employees have a set number of hours to work during the day, but they have different start and finish times.
- **Downsizing operation:** Reduction of a facility's production capacity accompanied with a reduction in the number of employees.
- **Adjusted sick day policy:** Employee benefits include a paid sick leave granted when an employee is unable to work because the employee is quarantined or isolated due to COVID-19, because of a bona fide need to care for an individual subject to quarantine or isolation, or to care for a child (under 18 years of age) whose school or child care provider is closed or unavailable for reasons related to COVID-19. (Definition is adapted from US Dept. of Labor "Families First Coronavirus Response Act: Employee Paid Leave Rights").
- **Spacing workers >6ft during production:** Keeping a space at least 6 feet between employees.
- **Cohorting employees:** Establishing groups of employees based on their risk of infection in the company, where each cohort remains as separated from the other cohorts as possible.

Q34. Have any of these **social distancing strategies** been applied in this facility/operation, **at any point** since the start of the COVID-19 pandemic?

| | Yes | Yes, but only partially/ temporarily | No |
|---|---|---|---|
| Installed physical barriers | ☐ | ☐ | ☐ |
| Staggered break times | ☐ | ☐ | ☐ |
| Staggered arrival/departure times (staggered shifts) | ☐ | ☐ | ☐ |
| Downsizing operation | ☐ | ☐ | ☐ |
| Adjusted sick day policy | ☐ | ☐ | ☐ |
| Spacing workers >6ft during production | ☐ | ☐ | ☐ |
| Cohorting employees | ☐ | ☐ | ☐ |

Q35. For one or more of these **social distancing** strategies, could you share **any reasons for adopting or not adopting** it, such as cost, compliance by workers, training requirement, effectiveness in reducing health risks, impact on production capacity, and/or lack of science-based information?

____________________________________________________________

____________________________________________________________

**<u>Block 5</u>:** Questions about different **employee biosafety** strategies for mitigation of COVID-19 in your facility/operation

***Definitions of employee biosafety mitigation strategies:***
• **Enhanced handwashing:** Implementation of a set of instructions for employees about when and how to wash hands that goes above and beyond instructions that were in place pre-COVID-19.
• **Alcohol-based hand rubs:** Implementation of a set of instructions for employees about when and how to use.
• **Face mask, face shields, and/or goggles:** Implementation of a set of instruction about how and when to use face masks, face shields and goggles. Face masks are often referred to as surgical masks or procedure masks. They cover nose and mouth and are secured under the chin, fit snugly against the side of the face and do not have gaps. Face shields are secondary protectors intended to protect the entire face against exposure. Goggles shield the eyes against the hazards.
• **Increased air ventilation rates:** Increase in the rate at which external air (fresh air) flows into the building.
• **Air cleaning/filtering**: destroying or removing hazards like viral particles from air.

Q36. Have any of these **employee biosafety strategies** been applied in this facility/operation, **at any point** since the start of the COVID-19 pandemic?

| | Yes | Yes, but only partially/ temporarily | No |
|---|---|---|---|
| Enhanced handwashing | ☐ | ☐ | ☐ |
| Alcohol-based hand rubs | ☐ | ☐ | ☐ |
| Face mask, face shields, goggles | ☐ | ☐ | ☐ |
| Increased ventilation rates | ☐ | ☐ | ☐ |
| Air cleaning/filtering | ☐ | ☐ | ☐ |

Q37. For one or more of these **employee biosafety strategies**, could you share **any reasons for adopting or not adopting** it, such as cost, compliance by workers, training requirement, effectiveness in reducing health risks, impact on production capacity, and/or lack of science-based information?

____________________________________________________________

____________________________________________________________

**<u>Block 6</u>:** Questions about different **surveillance** strategies for mitigation of COVID-19 in your facility/operation

***Definitions of surveillance mitigation strategies:***

• **Temperature screening and quarantine:** Screen for employees with temperature above 99.5°F (or other cut-off value) and keep identified employees away from workplace to determine whether they develop COVID-19 symptoms or test positive for the disease.

• **Test for infection and isolation:** Test employees for COVID-19 infection (viral test); Isolation: keep away from workplace an employee who is sick with COVID-19 or tested positive for COVID-19 without symptoms.

• **Contact tracing and quarantine:** Contact tracing is a process to identify individuals who may have been exposed to a person with COVID-19. Quarantine is the practice of separating individuals who have had close contact with someone with COVID-19 to determine whether they develop symptoms or test positive for the disease.

• **Return to work post recovery policy:** Any strategy implemented for employees returning to work following a COVID-19 infection based on symptoms or doctor's recommendation.

Q38. Have any of these **surveillance strategies** been applied in this facility/operation, **at any point** since the start of the COVID-19 pandemic?

| | Yes | Yes, but only partially/ temporarily | No |
|---|---|---|---|
| Temperature screening and quarantine | ☐ | ☐ | ☐ |
| Test for infection and isolation | ☐ | ☐ | ☐ |
| Contact tracing and quarantine | ☐ | ☐ | ☐ |
| Return to work post recovery policy | ☐ | ☐ | ☐ |

Q39. For one or more of these **surveillance strategies**, could you share **any reasons for adopting or not adopting** it, such as cost, compliance by workers, training requirement, effectiveness in reducing health risks, impact on production capacity, and/or lack of science-based information?

_______________________________________________

_______________________________________________

**<u>Block 7</u>:** Questions in this block ask about **any "Other" mitigation strategy** that was implemented or has been considered for implementation in your facility/operation, but we did not ask previously. If no "Other" strategies are applicable, skip to Block 8.

Q40. What is this **Other** mitigation strategy? Please specify.

____________________________________________________________

____________________________________________________________

Q41. Has this **Other** mitigation strategy been applied in this facility/operation, **at any point** since the start of the COVID-19 pandemic?

O Yes

O Yes, but only partially/ temporally

O No

Q42. Could you share **any reasons for adopting or not adopting** this **Other** strategy, such as cost, compliance by workers, training requirement, effectiveness in reducing health risks, impact on production capacity, and/or lack of science-based information?

____________________________________________________________

____________________________________________________________

**<u>Block 8</u>: Closing question** about your food production facility/operation

Q43. What was the **main reason** for your choice to describe conditions and COVID-19 mitigation in this particular food production facility/operation?
**(Response to this question is required)**

- O I am mostly familiar with this facility/operation
- O This is our unique facility/operation
- O This is our typical facility/operation
- O This facility/operation has been impacted greatly by COVID-19
- O This is our strategically important facility/operation
- O Other: ________

**Clicking the "Submit" button will SUBMIT your survey. If you would like to go back and check anything you answered, please do so now. You will not be able to access the survey after you submit it.**
