## Supplementary material for "Ongoing mitigation strategies and further needs of the United States food industry to control COVID-19 in the work environment": S1 Table

| **Q** | **Question**   - *Levels/Likert scale items* | **Question type** | **Analysis type^a^** | **Role in statistical analysis^b^** |
| --- | --- | --- | --- | --- |
| **Part 1: General questions about a participant’s industry sector** | | | | |
| 1^c^ | What industry sector are you in? (select all that apply)   - *Fresh produce; Dairy; Beef/Pork; Poultry; Other* | Multiple-choice | S | P |
| 2^c^ | Select your main role within your organization   - *C-suite; Regional manager; Facility manager; Research and development; Corporate food safety and quality; Other; Prefer not to answer* | Single-choice | S | P |
| 3 | Did COVID-19 have a significant impact on your industry sector?   - *Yes; No* | Single-choice | S | P |
| 4 | In which way(s) has COVID-19 significantly impacted your industry sector? (select all that apply)   - *Operations/production has been reduced/cut back; Operations/production has expanded; Implemented robotics, sensors, automation, and/or computer modeling; Management/corporate employees working remotely; Major changes in operational staffing control and protection protocols* | Multiple-choice | S | P |
| 5 | Regarding control of COVID-19 in your industry sector, how concerning are the items below?   - *Organizational awareness of the virus; Labor availability; Workers’ compliance with control measures; Workers’ abuse of control measures; Limited financial resources; Production capacity; Product quality; Supplier management; Customer expectations; Complex/ever-changing government regulations about COVID-19* | 5-point scale Likert question | S | Os |
| 6 | Regarding control of COVID-19 in your industry sector, are there any other concerns we should consider? | Open-ended | T | N/A |
| 7 | Regarding the labor force needed to maintain the production capacity in your industry sector during the COVID-19 pandemic, how challenging are the items below?   - *Access to number of workers needed; Access to workers with necessary skills; Sufficient housing for labor; Turnover in workforce; Need to train labor* | 5-point scale Likert question | S | Os |
| 8 | Regarding the labor force needed to maintain the production capacity in your industry sector during the COVID-19 pandemic, are there any other challenges we should consider? | Open-ended | T | N/A |
| 9^c^ | Regarding needs to successfully mitigate COVID-19 in your industry sector, how important are the items below?   - *More and better training; Training materials in more languages; Better technologies to assure social distancing; Better and cheaper testing technologies; Easier way to understand regulations; Better information on cost effectiveness of COVID-19 mitigation strategies* | 5-point scale Likert question | S | Op |
| 10 | Regarding needs to successfully mitigate COVID-19 in your industry sector, are there any other important needs we should consider? | Open-ended | T | N/A |
| 11^c^ | If computational modeling tools were available to predict which COVID-19 mitigation strategies would most likely be successful in a given facility/operation at a given time, how important would the model features below be for your industry sector?   - *Ability of the model to predict impact on production capacity; Ability of the model to predict initial and ongoing cost of implementation; Ability of the model to predict infection risk reduction; Ease of model use by company personnel; Ability to use the model confidentially; Ability to customize the model for use in a specific facility* | 5-point scale Likert question | S | Op |
| 12 | Regarding a potential modelling tool that could predict successful COVID-19 mitigation strategies, are there any other model features important for your industry sector we should consider? | Open-ended | None | N/A |
| 13^c^ | Regarding indicators of successful responses to COVID-19 in your industry sector, how important are the items below?   - *Workforce trained about COVID-19 risks and mitigation; Standard operating procedures/checklists are in place for mitigation of COVID-19 impacts; Established effective risk communication plan; Digital technologies utilized in planning of facility specific COVID-19 mitigation; Investment made into technologies that reduce vulnerability to a future pandemic or similar system wide disruption; Workforce related contingency plans updated to minimize COVID-19 related business interruptions* | 5-point scale Likert question | S | Op |
| 14 | Regarding attainable indicators of successful responses to COVID-19 in your industry sector, are there any other important indicators we should consider? | Open-ended | None | N/A |
| **Part 2: Conditions and COVID-19 controls in a facility or operation of participant’s choice** | | | | |
| 15^c^ | In what industry sector is this facility/operation?   - *Fresh produce; Dairy; Beef/Pork; Poultry; Other* | Single-choice | None | N/A |
| 16 | How does this facility/operation operate?   - *Year-round; Seasonally* | Single-choice | S | P |
| 17 | What are the approximate start and end dates for the production season(s) in this fresh produce facility/operation? | Open-ended | S | P |
| 18 | What role best describes this facility/operation?   - *Grower; Packing House; Processor; Grower and Field packer; Grower and Processor; Other* | Single-choice | S | P |
| 19 | Please select which part of your Grower and Processor operation will you describe in the remaining questions?   - *Grower operation; Processor facility* | Single-choice | S | P |
| 20 | What was the average number of employees in this facility/operation in 2019?   - *Less than 10; 10-49; 50-99; 100-249; 250-499; 500-999; 1000-2000; More than 2000; This facility/operation did not operate in 2019; Prefer not to answer* | Single-choice | S | P |
| 21 | What is the approximate proportion (%) of employees in this facility/operation that are between 50-69 years of age and 70 years old or older? | Open-ended | S | P |
| 22 | Does this facility/operation provide group temporary (seasonal) housing to any of your employees?   - *Yes; No* | Single-choice | S | P |
| 23 | Approximately what proportion (%) of employees in this facility/operation are provided with group temporary housing? | Open-ended | S | P |
| 24 | Does this facility/operation provide group transportation services (bus, truck, etc.) to employees to/from work?   - *Yes; No* | Single-choice | S | P |
| 25 | Approximately what proportion (%) of employees in this facility/operation are provided with group transportation to/from work? | Open-ended | S | P |
| 26^c^ | What is the largest percent reduction in the general production labor force that this facility/operation could withstand over a period of one week without reduction in the production capacity?   - *5%; 10%; 15%; 20%; 30%; 40%; 50%; Do not know* | Single-choice | S | Op |
| 27 | COVID-19 related work absences among workers performing different specialized job functions in this facility/operation present different levels of risk for a facility/operation shutdown. How would you describe the risk of a shutdown in this facility/operation due to work absences in each of the specialized job functions below?   - *Specialized production line functions (for example, operators for specialized equipment); Lab personnel; Quality control and assurance; Sanitation and cleaning; Supervisors; Engineering and/or maintenance crew* | 5-point scale Likert question | S | Os |
| 28 | Regarding the risk of a shutdown in this facility/operation due to work absences, are there any other specialized job functions we should consider? | Open-ended | None | N/A |
| 29 | Regarding potential sources of COVID-19 infection in this facility/operation, how concerning are the items below?   - *Employee housing conditions; Employee transportation conditions; Indoor common areas (for example, restrooms, breakrooms, personal protective equipment area, offices); Outdoor common areas; Common production tools/equipment; Production line; Activities in the local community* | 5-point scale Likert question | S | Os |
| 30 | Regarding potential sources of COVID-19 infection in this facility/operation, are there any other concerns we should consider? | Open-ended | None | N/A |
| 31 | If in the future the available labor force in this facility/operation is affected by a COVID-19 outbreak, what short/mid-term solutions should be considered to maintain production? (check all that apply)   - *Extend the number of work hours for remaining workers; Backfill with emergency personnel from third-party companies; Backfill by reorganizing personnel in the same facility* | Multiple-choice | S | P |
| 32 | To prevent labor shortage in this facility/operation due to a potential future COVID-19 outbreak, should capital investment into mechanization be considered as a long-term solution to maintain production?   - *Yes; No* | Single-choice | S | P |
| 33 | To maintain production in the event of labor shortage in this facility/operation due to a potential future COVID-19 outbreak, are there any other solutions we should consider? | Open-ended | T | N/A |
| 34 | Have any of these social distancing strategies been applied in this facility/operation, at any point since the start of the COVID-19 pandemic?   - *Installed physical barriers; Staggered break times; Staggered arrival/departure times (staggered shifts); Downsizing operation; Adjusted sick day policy; Spacing workers >6ft during production; Cohorting employees* | 3-point scale Likert question | S | Op |
| 35 | For one or more of these social distancing strategies, could you share any reasons for adopting or not adopting it, such as cost, compliance by workers, training requirement, effectiveness in reducing health risks, impact on production capacity, and/or lack of science-based information? | Open-ended | T | N/A |
| 36 | Have any of these employee biosafety strategies been applied in this facility/operation, at any point since the start of the COVID-19 pandemic?   - *Enhanced handwashing; Alcohol-based hand rubs; Face mask, face shields, goggles; Increased ventilation rates; Air cleaning/filtering* | 3-point scale Likert question | S | Op |
| 37 | For one or more of these employee biosafety strategies, could you share any reasons for adopting or not adopting it, such as cost, compliance by workers, training requirement, effectiveness in reducing health risks, impact on production capacity, and/or lack of science-based information? | Open-ended | T | N/A |
| 38 | Have any of these surveillance strategies been applied in this facility/operation, at any point since the start of the COVID-19 pandemic?   - *Temperature screening and quarantine; Test for infection and isolation; Contact tracing and quarantine; Return to work post recovery policy* | 3-point scale Likert question | S | Op |
| 39 | For one or more of these surveillance strategies, could you share any reasons for adopting or not adopting it, such as cost, compliance by workers, training requirement, effectiveness in reducing health risks, impact on production capacity, and/or lack of science-based information? | Open-ended | T | N/A |
| 40 | What is this Other mitigation strategy? Please specify. | Open-ended | None | N/A |
| 41 | Has this Other mitigation strategy been applied in this facility/operation, at any point since the start of the COVID-19 pandemic?   - *Yes; Yes, but only partially/ temporally; No* | 3-point scale Likert scale question | None | N/A |
| 42 | Could you share any reasons for adopting or not adopting this Other strategy, such as cost, compliance by workers, training requirement, effectiveness in reducing health risks, impact on production capacity, and/or lack of science-based information? | Open-ended | None | N/A |
| 43^c^ | What was the main reason for your choice to describe conditions and COVID-19 mitigation in this particular food production facility/operation?   - *I am mostly familiar with this facility/operation; This is our unique facility/operation; This is our typical facility/operation; This facility/operation has been impacted greatly by COVID-19; This is our strategically important facility/operation; Other* | Single-choice | S | P |

^a^ Analysis type: S=statistical analysis; T=thematic analysis; None=responses were examined but not systematically analyzed

^b^ Role in statistical analysis: Op=primary outcome; Os=secondary outcome; P=predictor; N/A=not applicable

^c^ Participants were required to answer the question to continue the needs assessment survey.
