## Supplementary material for "Ongoing mitigation strategies and further needs of the United States food industry to control COVID-19 in the work environment": S2 Table

| **Likert question** | **Corresponding open-ended question** (Number (N) responses) | **Theme** | **Code** | **Quote** | **Participant’s info** | |
| --- | --- | --- | --- | --- | --- | --- |
|  |  |  |  |  | Industry sector | ID |
| Q5 | Q6, Regarding control of COVID-19 in your industry sector, are there any other concerns we should consider? (N=19) | Access to COVID-19 preventative measures, guidance and information; difficulties in implementation of mitigation strategies | PPE shortage | Costs and the availability for PPE | Fresh produce | 40 |
|  |  |  | Vaccine access | Having vaccines available to our workforce. Being able to work with local agencies and providers to pre-arrange an on-site Covid-19 vaccination for our employees, much like flu vaccines are available. We encourage our employees to get vaccinated when they become eligible, however, it is so complex to try to arrange for an appointment, we are extremely concerned that our employees will not get vaccinated even when the vaccine is available to them. | Dairy | 11 |
|  |  |  | COVID-19 misinformation | Cultural attitudes/disinformation [about the] existence of the virus | Other (prepared food) | 51 |
|  |  |  | Difficulties in implementation of mitigation strategies | The bigger the food producer, the more likely to observe failures in preventative controls, including risk-reduction measures for Coronavirus SARS-II exposure and cross-transmission to fellow coworkers. Inspecting facilities for Food Safety issues also has exposed continual, chronic GMP and pre-requisite issues (including effective sanitation). | Other (fresh produce, dairy, beef/pork, and poultry) | 72 |
|  |  |  | Lack of COVID-19 guidance | Lack of industry specific guidance for food industry masking, face shields, and combo use. | Dairy | 71 |
|  |  | Employee fatigue, vaccine hesitancy, health and healthcare access | Employee fatigue | Employee (front-line production) fatigue. | Dairy | 29 |
|  |  |  | Vaccine hesitancy | Implementing vaccination programs with a somewhat resistant labor force | Fresh produce | 46 |
|  |  |  | Employee health | Mental health impact on managerial and office staff. In talking with industry colleagues, my experience is that many facilities have cut back on the staff responsible for ensuring that the facility is operating in an efficient and structured manner, while expanding the production capacity of the facility. It leaves many of these employees in a position where they are stretched thin and feel overwhelmed. Conversely, many facilities are putting policies in place to improve the quality of life for hourly workers to try to attract and keep workers in an environment where it is hard to find new workers at wage rates that previously attracted workers. | Other (co-packaging of shelf stable products) | 75 |
|  |  |  | Employee healthcare access | Transient work populations working in remote areas with limited public health resources | Other (seafood) | 76 |
|  |  | Supply chain disruption and management of contractor expectations | Supply chain disruption | Ability of suppliers upstream of us. | Dairy | 56 |
|  |  |  | Management of contractor expectations | Contractor control measures are difficult to manage. | Dairy | 16 |
| Q7 | Q8, Regarding the labor force needed to maintain the production capacity in your industry sector during the COVID-19 pandemic, are there any other challenges we should consider? (N=20) | Government benefits and regulations | Government benefits for unemployed | Easily obtainable government handouts have a negative effect on those willing to participate in the labor market | Dairy | 9 |
|  |  |  | Changing government regulations | Government directives and laws regarding labor, time off, and pay rules and the changes in these made managing government special rules a full time job, rather than managing the pandemic we were managing the labor law compliance tasks. | Fresh produce | 45 |
|  |  | Downside of COVID-19 mitigation strategies | Downside of mitigations | Because of social distancing, infrastructure is not enough to keep the amount of people we need to reach our production goals | Fresh produce | 42 |
|  |  |  |  | Housing and transportation of workforce to enable safe social distancing during the pandemic  and contact tracing reducing large portions of the normal workforce in quick succession | Fresh produce | 46 |
|  |  | Labor availability, needs, expectations and behavior | Inability to hire skilled labor | Finding a qualified temporary work force. | Dairy | 15 |
|  |  |  | Employee behavior | Employees getting, spreading, being exposed to the virus. | Other (food services) | 39 |
|  |  |  | Reliance on critical employees | When someone is out of the workplace sick it is incredibly difficult to manage for small businesses | Other (food processing) | 58 |
|  |  |  | Bias in hiring minorities | Recent events in Minneapolis have shown the difficulties certain minorities group face and this is pushing the industry to reevaluate the recruiting process to ensure unbiased hiring practices. | Other (spirits) | 62 |
|  |  |  | Unacceptable employee expectations | Expectation of pay. In Northern NJ, average pay expectation has increased as Amazon offers close to 20 an hour for minimally skilled labor. Grocery stores are also offering close to 15 for the same. There are many food facilities who were previously between 11 (state minimum) and 15 an hour for unskilled labor. Customers aren't always willing to accept price increases. So it is a balance of splitting increased labor costs with customers, which makes thin margins thinner. | Other (co-packaging of shelf stable products) | 75 |
|  |  |  | Employee support services | Transportation. Childcare | Fresh produce | 65 |
|  |  |  | Employee language barriers | Multi language barriers | Other (seafood) | 76 |
| Q9 | Q10, Regarding needs to successfully mitigate COVID-19 in your industry sector, are there any other important needs we should consider? (N=8) | Consumer education | Consumer education | In regards to food industry, zero cases food to people, people to food must be communicated to public | Fresh produce | 60 |
|  |  | Technology to improve infection prevention, time efficiency and internet access | Technology to reduce infection exposure | More technology to reduce exposure and maximize time efficiency | Fresh produce | 42 |
|  |  |  | Technology to improve time efficiency | More technology to reduce exposure and maximize time efficiency | Fresh produce | 42 |
|  |  |  | High-speed Internet | Remote areas with little to no internet makes training and communication difficult | Other (seafood) | 76 |
|  |  | Cost-effective mitigation strategies, harmonized guidance and prioritized vaccination of food industry workers | Harmonized guidance about COVID-19 control | Consider that during the CV-19 pandemic businesses have received directives and policy recommendations and requirements from: Federal, state, and local governments, industry trade associations, our insurance company, food safety consultants, legal teams, auditors. All with different recommendations…none of them were right...ridiculous We've turned the pandemic into a new industry. | Fresh produce | 45 |
|  |  |  | Assistance to small-to-medium-sized businesses | Better support for small to medium sized farms/companies, including expansion of extension resources. | Other (unspecified^a^) | 50 |
|  |  |  | Cost-effective mitigation strategies | No [return of investment] ROI assessments have been made in effective mitigation techniques; their have been no incentives to implement mitigation techniques; no enforcement on simple requirements led many not to implement effective measurements | Other (spirits) | 62 |
|  |  |  | Prioritized food worker vaccination | Vaccination of "front-line" clinical workers needs to also be re-prioritized to include all of the front-line food handlers. | Other (fresh produce, dairy, beef/pork, and poultry) | 72 |
| Q32 | Q33, To maintain production in the event of labor shortage in this facility/operation due to a potential future COVID-19 outbreak, are there any other solutions we should consider? (N=9) | Employee benefits and training | Employee benefits | If extending the number of work hours for remaining workers, compensate with double time for hours worked past 40, Start OT after scheduled hours worked daily instead of after 40 hours | Dairy | 8 |
|  |  | Industry collaboration, production adjustment and infrastructure changes | Collaboration across industry | Possibly organize and facilitate an exchange program amongst dairy processing companies to support one another with qualified workers. | Dairy | 9 |
|  |  |  | Adjust production | Reduction of the complexity of [Stock Keeping Unit] SKU offerings to maximize efficient production and maximize limited offerings at higher volumes to all customers | Fresh produce | 46 |
|  |  |  | Infrastructure changes | Enlarging employee common areas. |  | 56 |
| Q34 | Q35, For one or more of these social distancing strategies, could you share any reasons for not adopting it, such as cost, compliance by workers, training requirement, effectiveness in reducing health risks, impact on production capacity, and/or lack of science-based information? (N=12) | Infrastructure, productivity or Union imposed constraints | Infeasible due to infrastructure limitations | Space considerations have been implemented but there are occasions which plant design does not allow 100% effectiveness. We mitigate by changing production schedules and staggering production to limit number of employees in close contact while on the floor | Fresh produce | 46 |
|  |  |  | Negative impact on productivity | If the plant does not run near full production, we are out of business. | Dairy | 56 |
|  |  |  |  | We are unable to downsize due to connection with farmers. | Dairy | 9 |
|  |  |  | Union imposed constraints | Cohorting due to union contract constraints. | Dairy | 30 |
|  |  |  | No improvements applicable | We do not have enough employees to make staggering shift a possible solution | Other (food processing) | 58 |
|  |  | Lack of concern or need | Lack of concern | All employees are very diligent in following the rules, so We do not feel we need to worry as much | Other (manufacturing shelf-stable foods) | 63 |
|  |  |  | No improvements required or applicable | We are small and family the of the four have been vaccinated. | Fresh produce | 59 |
| Q36 | Q37, For one or more of these biosafety strategies, could you share any reasons for not adopting it, such as cost, compliance by workers, training requirement, effectiveness in reducing health risks, impact on production capacity, and/or lack of science-based information? (N=10) | Lack of funds, supplies or information | Unavailable supplies | Availability of supplies at the onset but now you need to monitor the reliability of the products for compliance. | Dairy | 4 |
|  |  |  | Cost of implementation | We felt the cost of a new or upgraded ventilation system was not justified with all of the other precautions already in place. | Other (manufacturing shelf-stable foods) | 63 |
|  |  |  | Lack of information on effectiveness | Leadership not willing to make decisions, no CDC guidance on benefits for doing more than normally done. Mask usage yes, face shields no due to no CDC guidance for combo. No advisement on managing ventilation and filtering | Dairy | 71 |
|  |  | Infrastructure constraints | Infeasible due to infrastructure limitations | Increased ventilation is applied when possible but the nature of our production requires controlled environment inside the building. Air filtering is not as practical in a short term implementation -we use positive air in high density work areas | Fresh produce | 46 |
|  |  | Lack of need | No improvements required or applicable | Plant air is already highly filtered with turnover every 20 minutes, so no change there. | Dairy | 56 |
| Q38 | Q39, For one or more of these surveillance strategies, could you share any reasons for not adopting it, such as cost, compliance by workers, training requirement, effectiveness in reducing health risks, impact on production capacity, and/or lack of science-based information? (N=10) | Lack of concern or need | Lack of concern | We all have been and continue to be very careful and healthy. | Other (manufacturing shelf-stable foods) | 63 |
|  |  |  | No improvements required or applicable | Good community testing access so no reason to bring on site | Dairy | 30 |
|  |  | Increases worker absences | Increases work absences | No funding available. | Dairy | 56 |

^a^Academic affiliated with the food industry.
