## Supplementary material for "Ongoing mitigation strategies and further needs of the United States food industry to control COVID-19 in the work environment": S3 Table

| **Likert question (Q)^@^**   - Item | **Predictor** | **Statistical test** | **Level** | **Median^*^/correlation coefficient** | **IQR^*^** | ***p*-value** |
| --- | --- | --- | --- | --- | --- | --- |
| Q5, Regarding control of COVID-19 in your industry sector, how concerning are the items below? |  |  |  |  |  |  |
| - Limited financial resources | Q4, COVID-19 impact on industry sector: Operations/production has expanded | MW-U | No | 3 | 2-4 | <0.001 |
|  |  |  | Yes | 1 | 1-2 |  |
| - Limited financial resources | Q4, COVID-19 impact on industry sector: Operations/production has been reduced/cut back | MW-U | No | 2 | 1-3 | 0.02 |
|  |  |  | Yes | 3 | 2-4 |  |
| - Limited financial resources | Q4, COVID-19 impact on industry sector: Implemented robotics, sensors, automation, and/or computer modeling | MW-U | No | 2 | 2-3.25 | 0.04 |
|  |  |  | Yes | 4 | 3-4 |  |
| - Limited financial resources | Q31, Short mid solutions to maintain production: Extend the number of work hours for remaining workers | MW-U | No | 4 | 2-4 | 0.03 |
|  |  |  | Yes | 2 | 1.25-3 |  |
| - Limited financial resources | Q22, Temporary housing provided to employees | MW-U | No | 2 | 2-3 | 0.02 |
|  |  |  | Yes | 4 | 3-4 |  |
| - Complex/ever-changing government regulations about COVID-19 | Q4, COVID-19 impact on industry sector: Operations/production has expanded | MW-U | No | 4 | 3-5 | 0.02 |
|  |  |  | Yes | 3 | 3-4 |  |
| - Complex/ever-changing government regulations about COVID-19 | Q4, COVID-19 impact on industry sector: Major changes in operational staffing control and protection protocols | MW-U | No | 3 | 3-4 | 0.04 |
|  |  |  | Yes | 4 | 3-5 |  |
| - Workers’ compliance with control measures | Q23, Proportion (%) of employees with group temporary housing | SC | N/A^#^ | -0.81 | N/A | 0.03 |
| - Workers’ compliance with control measures | Q25, Proportion (%) of employees with group transportation | SC | N/A | 0.85 | N/A | 0.02 |
| - Customer expectations | Q4, COVID-19 impact on industry sector: Operations/production has expanded | MW-U | No | 3 | 2-4 | 0.03 |
|  |  |  | Yes | 2 | 2-3 |  |
| - Customer expectations | Q4, COVID-19 impact on industry sector: Major changes in operational staffing control and protection protocols | MW-U | No | 2 | 1-3 | 0.005 |
|  |  |  | Yes | 3 | 2-4 |  |
| - Customer expectations | Q31, Backfill by reorganizing personnel in the same facility | MW-U | No | 2 | 1.75-3 | 0.02 |
|  |  |  | Yes | 3 | 2-4 |  |
| - Organizational awareness of the virus | Q25, Proportion (%) of employees with group transportation | SC | N/A | 0.85 | N/A | 0.02 |
| Q7, Regarding the labor force needed to maintain the production capacity in your industry sector during the COVID-19 pandemic, how challenging are the items below? |  |  |  |  |  |  |
| - Access to workers with necessary skills | Q4, COVID-19 impact on industry sector: Major changes in operational staffing control and protection protocols | MW-U | No | 2.5 | 2-3.25 | 0.04 |
|  |  |  | Yes | 4 | 3-3.45 |  |
| - Sufficient housing for labor | Q4, COVID-19 impact on industry sector: Implemented robotics, sensors, automation, and/or computer modeling | MW-U | No | 2 | 1-3 | 0.001 |
|  |  |  | Yes | 4 | 4-4 |  |
| - Sufficient housing for labor | Q22, Temporary housing provided to employees | MW-U | No | 2 | 1-3 | 0.004 |
|  |  |  | Yes | 4 | 3-5 |  |
| - Sufficient housing for labor | Q21, Proportion (%) of employees 70 years or older^%^ | SC | N/A | 0.40 | N/A | 0.004 |
| Q9, Regarding needs to successfully mitigate COVID-19 in your industry sector, how important are the items below? |  |  |  |  |  |  |
| - Better information on cost effectiveness of COVID-19 mitigation strategies | Q4, COVID-19 impact on industry sector: Operations/production has expanded | MW-U | No | 4 | 3-4 | 0.005 |
|  |  |  | Yes | 3 | 2-3.5 |  |
| - Training materials in more languages | Q4, COVID-19 impact on industry sector: Operations/production has expanded | MW-U | No | 3 | 2-4 | 0.002 |
|  |  |  | Yes | 1 | 1-2.5 |  |
| - Training materials in more languages | Q4, COVID-19 impact on industry sector: Operations/production has been reduced/cut back | MW-U | No | 2 | 1-2 | <0.001 |
|  |  |  | Yes | 4 | 2-4 |  |
| - Training materials in more languages | Q22, Temporary housing provided to employees | MW-U | No | 2 | 1-4 | 0.02 |
|  |  |  | Yes | 5 | 3-5 |  |
| - Better and cheaper testing technologies | Q4, COVID-19 impact on industry sector: Operations/production has expanded | MW-U | No | 4 | 3-5 | 0.02 |
|  |  |  | Yes | 3 | 2-4 |  |
| - Better and cheaper testing technologies | Q4, COVID-19 impact on industry sector: Operations/production has been reduced/cut back | MW-U | No | 3 | 2-4 | 0.002 |
|  |  |  | Yes | 4 | 3-5 |  |
| - Better and cheaper testing technologies | Q22, Temporary housing provided to employees | MW-U | No | 3 | 2-4 | 0.03 |
|  |  |  | Yes | 5 | 3-5 |  |
| - More and better training | Q4, COVID-19 impact on industry sector: Operations/production has been reduced/cut back | MW-U | No | 3 | 2-4 | 0.02 |
|  |  |  | Yes | 4 | 3-5 |  |
| - More and better training | Q24, Group transportation provided to employees | MW-U | No | 3 | 2-4 | 0.02 |
|  |  |  | Yes | 4 | 4-5 |  |
| - Easier way to understand regulations | Q4, COVID-19 impact on industry sector: Operations/production has expanded | MW-U | No | 4 | 3-5 | 0.03 |
|  |  |  | Yes | 3 | 2-4 |  |
| - Easier way to understand regulations | Q4, COVID-19 impact on industry sector: Management/corporate employees working remotely | MW-U | No | 2.5 | 2-3.75 | 0.008 |
|  |  |  | Yes | 4 | 3-5 |  |
| - Better technologies to assure social distancing | Q25, Proportion (%) of employees with group transportation | SC | N/A | 0.78 | N/A | 0.04 |
| - Better technologies to assure social distancing | Q4, COVID-19 impact on industry sector: Operations/production has been reduced/cut back | MW-U | No | 3 | 2-3 | 0.01 |
|  |  |  | Yes | 4 | 2-4 |  |
| Q11, If computational modeling tools were available to predict which COVID-19 mitigation strategies would most likely be successful in a given facility/operation at a given time, how important would the model features below be for your industry sector? |  |  |  |  |  |  |
| - Ease of model use by company personnel | Q4, COVID-19 impact on industry sector: Operations/production has expanded | MW-U | No | 4 | 3-5 | 0.003 |
|  |  |  | Yes | 3 | 3-4 |  |
| - Ability to customize the model for use in a specific facility | Q4, COVID-19 impact on industry sector: Operations/production has expanded | MW-U | No | 4 | 4-5 | 0.007 |
|  |  |  | Yes | 3.5 | 3-4 |  |
| - Ability of the model to predict infection risk reduction | Q4, COVID-19 impact on industry sector: Operations/production has expanded | MW-U | No | 4 | 3-5 | 0.03 |
|  |  |  | Yes | 3.5 | 3-4 |  |
| - Ability of the model to predict initial and ongoing cost of implementation | Q4, COVID-19 impact on industry sector: Operations/production has been reduced/cut back | MW-U | No | 3 | 2.75-4 | 0.02 |
|  |  |  | Yes | 4 | 3-5 |  |
| Q13, Regarding indicators of successful responses to COVID-19 in your industry sector, how important are the items below? |  |  |  |  |  |  |
| - Digital technologies utilized in planning of facility specific COVID-19 mitigation | Q4, COVID-19 impact on industry sector: Operations/production has expanded | MW-U | No | 3.5 | 3-4 | 0.02 |
|  |  |  | Yes | 3 | 2-3 |  |
| - Digital technologies utilized in planning of facility specific COVID-19 mitigation | Q4, COVID-19 impact on industry sector: Operations/production has been reduced/cut back | MW-U | No | 3 | 2-3 | 0.04 |
|  |  |  | Yes | 4 | 3-4 |  |
| - Digital technologies utilized in planning of facility specific COVID-19 mitigation | Q4, COVID-19 impact on industry sector: Implemented robotics, sensors, automation, and/or computer | MW-U | No | 3 | 2-4 | 0.05 |
|  |  |  | Yes | 4 | 4-4 |  |
| - Digital technologies utilized in planning of facility specific COVID-19 mitigation | Q4, COVID-19 impact on industry sector: Major changes in operational staffing control and protection protocols | MW-U | No | 2 | 2-3 | 0.04 |
|  |  |  | Yes | 3 | 3-4 |  |
| - Digital technologies utilized in planning of facility specific COVID-19 mitigation | Q25, Proportion (%) of employees with group transportation | SC | N/A | 0.88 |  | 0.009 |
| - Digital technologies utilized in planning of facility specific COVID-19 mitigation | Q4, COVID-19 impact on industry sector: Operations/production has expanded | MW-U | No | 4 | 3-5 | 0.03 |
|  |  |  | Yes | 3 | 2-4 |  |
| - Workforce trained about COVID-19 risks and mitigation | Q4, COVID-19 impact on industry sector: Major changes in operational staffing control and protection protocols | MW-U | No | 4 | 3-4 | 0.03 |
|  |  |  | Yes | 4 | 4-5 |  |
| - Standard operating procedures/checklists are in place for mitigation of COVID-19 impacts | Q4, COVID-19 impact on industry sector: Major changes in operational staffing control and protection protocols | MW-U | No | 4 | 3-4 | 0.03 |
|  |  |  | Yes | 4 | 4-5 |  |
| - Established effective risk communication plan | Q4, COVID-19 impact on industry sector: Major changes in operational staffing control and protection protocols | MW-U | No | 3 | 3-4 | 0.007 |
|  |  |  | Yes | 4 | 4-5 |  |
| - Workforce related contingency plans updated to minimize COVID-19 related business interruptions | Q31, Backfill by reorganizing personnel in the same facility | MW-U | No | 3 | 2.75-4 | 0.02 |
|  |  |  | Yes | 4 | 3.75-5 |  |
| Q27, COVID-19 related work absences among workers performing different specialized job functions in this facility/operation present different levels of risk for a facility/operation shutdown. How would you describe the risk of a shutdown in this facility/operation due to work absences in each of the specialized job functions below? |  |  |  |  |  |  |
| - Sanitation and cleaning | Q4, COVID-19 impact on industry sector: Management/corporate employees working remotely | MW-U | No | 3 | 2-3.75 | 0.01 |
|  |  |  | Yes | 4 | 3-5 |  |
| - Lab personnel | Q4, COVID-19 impact on industry sector: Management/corporate employees working remotely | MW-U | No | 2.5 | 2-3 | 0.03 |
|  |  |  | Yes | 3 | 3-4 |  |
| - Lab personnel | Q25, Proportion (%) of employees with group transportation | SC | N/A | -0.90 | N/A | 0.02 |
| - Supervisors | Q4, COVID-19 impact on industry sector: Management/corporate employees working remotely | MW-U | No | 3 | 2-3 | 0.02 |
|  |  |  | Yes | 3 | 3-4 |  |
| - Engineering and/or maintenance crew | Q21, Proportion (%) of employees from 50 to 69 years^a^ | SC | N/A | 0.27 | N/A | 0.04 |
| Q29, Regarding potential sources of COVID-19 infection in this facility/operation, how concerning are the items below? |  |  |  |  |  |  |
| - Activities in the local community | Q4, COVID-19 impact on industry sector: Major changes in operational staffing control and protection protocols | MW-U | No | 2 | 1-3 | <0.001 |
|  |  |  | Yes | 4 | 3-5 |  |
| - Indoor common areas (for example, restrooms, breakrooms, personal protective equipment area, offices) | Q22, Temporary housing provided to employees | MW-U | No | 3 | 2-4 | 0.02 |
|  |  |  | Yes | 4 | 3-5 |  |
| - Common production tools/equipment | Q31, Backfill by reorganizing personnel in the same facility | MW-U | No | 2 | 1.75-3 | 0.03 |
|  |  |  | Yes | 3 | 2-4 |  |
| - Common production tools/equipment | Q21, Proportion (%) of employees from 70 years or older^c^ | SC | N/A | 0.32 | N/A | 0.02 |
| - Production line | Q4, COVID-19 impact on industry sector: Major changes in operational staffing control and protection protocols | MW-U | No | 2 | 1.5-2.5 | 0.01 |
|  |  |  | Yes | 3 | 2-4 |  |
| - Production line | Q24, Group transportation provided to employees | MW-U | No | 3 | 2-3 | 0.04 |
|  |  |  | Yes | 4 | 2.75-4 |  |
| - Employee housing conditions | Q4, COVID-19 impact on industry sector: Operations/production has expanded | MW-U | No | 3 | 1-4 | 0.04 |
|  |  |  | Yes | 1 | 1-2 |  |
| - Employee housing conditions | Q4, COVID-19 impact on industry sector: Major changes in operational staffing control and protection protocols | MW-U | No | 1 | 1-2 | 0.03 |
|  |  |  | Yes | 2.5 | 1-4 |  |
| - Employee housing conditions | Q22, Temporary housing provided to employees | MW-U | No | 1 | 1-3 | <0.001 |
|  |  |  | Yes | 4 | 3-5 |  |
| - Employee housing conditions | Q24, Group transportation provided to employees | MW-U | No | 1.5 | 1-3 | 0.02 |
|  |  |  | Yes | 3.5 | 2.75-3.5 |  |
| - Employee transportation conditions | Q4, COVID-19 impact on industry sector: Management/corporate employees working remotely | MW-U | No | 1 | 1-2.5 | 0.05 |
|  |  |  | Yes | 2 | 1-4 |  |
| - Employee transportation conditions | Q4, COVID-19 impact on industry sector: Major changes in operational staffing control and protection protocols | MW-U | No | 1 | 1-1 | 0.002 |
|  |  |  | Yes | 2.5 | 1-4 |  |
| - Employee transportation conditions | Q22, Temporary housing provided to employees | MW-U | No | 1.5 | 1-3 | 0.002 |
|  |  |  | Yes | 4 | 3-5 |  |
| - Employee transportation conditions | Q24, Group transportation provided to employees | MW-U | No | 1 | 1-3 | 0.007 |
|  |  |  | Yes | 3.5 | 2.75-4.25 |  |
| - Outdoor common areas | Q4, COVID-19 impact on industry sector: Major changes in operational staffing control and protection protocols | MW-U | No | 1 | 1-1.5 | 0.01 |
|  |  |  | Yes | 2 | 1-3 |  |
| - Outdoor common areas | Q21, Proportion (%) of employees from 70 years or older^c^ | SC | N/A | 0.27 | N/A | 0.05 |
| - Outdoor common areas | Q24, Group transportation provided to employees | MW-U | No | 1 | 1-2 | 0.005 |
|  |  |  | Yes | 3 | 2-4 |  |
| Q34, Have any of these social distancing strategies been applied in this facility/operation, at any point since the start of the COVID-19 pandemic? |  |  |  |  |  |  |
| - Downsizing operation | Q4, COVID-19 impact on industry sector: Operations/production has expanded | MW-U | No | 0 | 0-0.5 | 0.004 |
|  |  |  | Yes | 0 | 0-0 |  |
| - Downsizing operation | Q4, COVID-19 impact on industry sector: Operations/production has been reduced/cut back | MW-U | No | 0 | 0-0 | 0.002 |
|  |  |  | Yes | 0 | 0-0.5 |  |
| - Downsizing operation | Q22, Temporary housing provided to employees | MW-U | No | 0 | 0-0 | 0.003 |
|  |  |  | Yes | 0.5 | 0-0.5 |  |
| - Downsizing operation | Q21, Proportion (%) of employees from 70 years or older^c^ | SC | N/A | 0.31 | N/A | 0.02 |
| - Downsizing operation | Q25, Proportion (%) of employees with group transportation | SC | N/A | 0.88 | N/A | 0.009 |
| - Adjusted sick day policy | Q23, Proportion (%) of employees with group temporary housing | SC | N/A | -0.76 | N/A | 0.05 |
| - Staggered arrival/departure times (staggered shifts) | Q4, COVID-19 impact on industry sector: Major changes in operational staffing control and protection protocols | MW-U | No | 0 | 0-0.75 | 0.03 |
|  |  |  | Yes | 1 | 0.125-1 |  |
| - Cohorting employees | Q4, COVID-19 impact on industry sector: Management/corporate employees working remotely | MW-U | No | 0 | 0-0.375 | 0.02 |
|  |  |  | Yes | 0.5 | 0-1 |  |
| - Cohorting employees | Q31, Short mid solutions to maintain production: Backfill with emergency personnel from third-party companies | MW-U | No | 0 | 0-0.5 | 0.02 |
|  |  |  | Yes | 0.5 | 0-1 |  |
| - Staggered break times | Q4, COVID-19 impact on industry sector: Operations/production has expanded | MW-U | No | 1 | 0.5-1 | 0.05 |
|  |  |  | Yes | 1 | 1-1 |  |
| - Staggered break times | Q4, COVID-19 impact on industry sector: Management/corporate employees working remotely | MW-U | No | 0.75 | 0-1 | 0.03 |
|  |  |  | Yes | 1 | 1-1 |  |
| - Installed physical barriers | Q4, COVID-19 impact on industry sector: Major changes in operational staffing control and protection protocols | MW-U | No | 0.5 | 0-0.5 | 0.006 |
|  |  |  | Yes | 1 | 0.5-1 |  |
| Q36, Have any of these employee biosafety strategies been applied in this facility/operation, at any point since the start of the COVID-19 pandemic? |  |  |  |  |  |  |
| - Increased ventilation rates | Q31, Backfill by reorganizing personnel in the same facility | MW-U | No | 0 | 0-0.5 | 0.05 |
|  |  |  | Yes | 0.5 | 0-1 |  |
| - Air cleaning/filtering | Q4, COVID-19 impact on industry sector: Implemented robotics, sensors, automation, and/or computer | MW-U | No | 0.5 | 0-1 | 0.03 |
|  |  |  | Yes | 1 | 1-1 |  |
| - Air cleaning/filtering | Q31, Backfill by reorganizing personnel in the same facility | MW-U | No | 0 | 0-0.625 | 0.04 |
|  |  |  | Yes | 0.5 | 0-1 |  |
| - Enhanced handwashing | Q25, Proportion (%) of employees with group transportation | SC | N/A | 0.79 | N/A | 0.03 |
| - Alcohol-based hand rubs | Q25, Proportion (%) of employees with group transportation | SC | N/A | 0.79 | N/A | 0.03 |
| Q38, Have any of these surveillance strategies been applied in this facility/operation, at any point since the start of the COVID-19 pandemic? |  |  |  |  |  |  |
| - Contact tracing and quarantine | Q4, COVID-19 impact on industry sector: Operations/production has expanded | MW-U | No | 1 | 0.5-1 | 0.01 |
|  |  |  | Yes | 1 | 1-1 |  |
| - Return to work post recovery policy | Q4, COVID-19 impact on industry sector: Operations/production has expanded | MW-U | No | 1 | 0.5-1 | 0.008 |
|  |  |  | Yes | 1 | 1-1 |  |
| - Return to work post recovery policy | Q4, COVID-19 impact on industry sector: Management/corporate employees working remotely | MW-U | No | 0.5 | 0-1 | 0.01 |
|  |  |  | Yes | 1 | 1-1 |  |
| - Test for infection and isolation | Q24, Group transportation provided to employees | MW-U | No | 0.5 | 0-1 | 0.01 |
|  |  |  | Yes | 1 | 1-1 |  |

^@^ Likert question (Q) number in the needs assessment survey.

^*^ Median and interquartile range (IQR) calculated for the interval (1-5 in Q5, Q9, Q13, and Q29) and numeric values (0, 0.5 or 1 in Q34, Q36, and Q38) that were assigned to the Likert scale responses in each Likert item are included for comparison across facility/operation size and industry sector levels. For Q5 and Q29, values 1 to 5 represented “Not at all concerning” to “Extremely concerning”; while for Q9 and Q13, values 1 to 5 represented “Not at all important” to “Extremely important”. For Q34, Q36, and Q38, the value 0 represented “No” (not implemented), 0.5 represented “Yes, but only partially/temporarily, and 1 represented “Yes” (implemented).

^#^ N/A=not applicable

^%^ The original question asked about ‘50-69 years of age’ and ‘70 years or older’ but were separated here to more clearly indicate their values.
